# Transdiagnostic Neurobehavioral Gradients and Environmental Interactions in Youth with Major Psychiatric Disorders

**DOI:** 10.64898/2026.02.04.26345162

**Authors:** Xiaofen Zong, Yi Ye, Jinxin He, Kaitong Ma, Mang Ye, Tao Yao, Siwei Li, He Li, Ge Song, Yinshan Wang, Bing Xiang Yang, Mengyao Feng, Qi Wen, Jie Yao, Li Dong, Xia Sun, Yuanyuan Zhang, Maolin Hu, Xinian Zuo, Lifespan Brain Chart Consortium (LBCC), Xujun Duan, Li Zhang

## Abstract

Major psychiatric disorders typically emerge in youth and exhibit shared and disorder-specific behavioral phenotypes and neuroanatomical alterations, yet the transdiagnostic neurobehavioral gradients and environmental interactions contributing to this heterogeneity remain poorly understood. Here, we present a transdiagnostic cohort of 1,755 youths aged 10-24 years, including 1,040 patients with bipolar disorder (BD), major depressive disorder (MDD), or schizophrenia spectrum disorder, and 715 healthy controls. Individualized gray matter volume (GMV) were quantified relative to population-based norms and integrated with behavioral phenotypes and environmental exposures. We identified transdiagnostic severity gradients across emotional and non-emotional symptoms, cognition, and personality traits, alongside widespread negative GMV deviations and diagnosis-specific effects in the pars opercularis and posterior cingulate, key hubs of the action-mode network orchestrating goal-directed functions. Two brain-behavior modes were identified: a cognitive mode linking posterior cortical variation with processing speed and an emotional mode associating prefrontal regions and the paracentral lobule with self-injurious behaviors. Further analyses indicated that adverse social environments were indirectly associated with brain structural deviations through behavioral pathways in BD and MDD, whereas air pollution (PM_2.5_) specifically moderated brain-behavior relationships in MDD. Together, these findings elucidate transdiagnostic neurobehavioral gradients across youth psychiatric disorders, with environmental exposures differentially embedded within neurobehavioral systems.

## Introduction

Mental disorders affect approximately one in seven individuals worldwide^1^ and typically emerge early in life, with 75% manifesting prior to age 24^2^. Schizophrenia spectrum disorders (SSD), bipolar disorder (BD), and major depressive disorder (MDD) represent the most prevalent major psychiatric disorders (MPDs). Youth with these conditions face profound lifelong disability and socioeconomic burden, driven by persistent symptomatology, compounded healthcare needs, and cumulative losses in education and employment^3–5^. MPDs have been proposed as neurodevelopmental disorders^6–8^, and are commonly characterized by behavioral phenotypes that include emotional symptoms (e.g., depression, anxiety, mania, and hypomania), psychotic symptoms, and cognitive deficits. Understanding the neuropathological mechanisms underlying these phenotypes during youth is therefore a major public health priority.

Psychiatric diagnosis has traditionally relied on categorical classification systems, such as the Diagnostic and Statistical Manual of Mental Disorders (DSM). While these systems remain clinically valuable, converging evidence from neuroimaging, genetics, and psychopathology increasingly challenges the notion that DSM-defined disorders represent discrete biological entities. Instead, MPDs exhibit substantial overlap in symptoms, cognitive deficits, and neural alterations^9–14^, suggesting that core neurobehavioral dimensions may transcend diagnostic boundaries. This recognition has motivated the development of transdiagnostic frameworks, such as the NIMH Research Domain Criteria (RDoC)^15^, which aim to capture both shared and disorder-specific mechanisms across continuous dimensions of psychopathology.

Neuroimaging studies have reported structural abnormalities across multiple MPDs, particularly in frontal, cingulate, and posterior cortical regions^16–18^. However, the consistency and graded severity of these alterations remain unclear across the literature. A major methodological limitation is that most studies rely on disorder-specific samples without a common normative reference^11,19,20^, hindering direct cross-disorder comparisons and obscuring potential transdiagnostic gradients of brain structure. Brain chart normative modeling addresses this limitation by benchmarking individual neuroanatomical measures against large-scale population-derived reference trajectories^21,22^. Analogous to pediatric growth charts, this approach enables quantification of individual deviations from normative neurodevelopment and supports systematic comparisons across diagnostic groups. Although normative modeling has recently been applied to psychiatric populations^23,24^, its potential to characterize neuroanatomical deviations and elucidate brain-behavior relationships in youth with MPDs remains largely unexplored.

Beyond intrinsic neural alterations, environmental exposures represent critical external influences on mental health. Social environments, encompassing both supportive factors (e.g., social support^25,26^, parental warmth^27^) and adverse factors (e.g., childhood trauma^28,29^, negative parenting^30^), and natural environments, including green space exposure^31,32^ and fine particulate matter (PM_2.5_)^33^, have been linked to symptom severity and cognitive outcomes. Emerging evidence further indicates that behavioral phenotypes are closely coupled with neural alterations^19,34–37^. Together, these findings suggest that behavior may serve as a potential intermediate pathway through which environmental exposures become biologically embedded in the developing brain. For example, behavioral problems have been shown to mediate the association between elevated family conflict and reduced cortical surface area^38^. Importantly, environmental influences may extend beyond mediating pathways to moderate the coupling between brain and behavioral phenotypes. Preliminary evidence suggests that environmental contexts can amplify or attenuate the strength and direction of brain-behavior associations^39^. However, a comprehensive examination of environment-behavior-brain triadic interactions in youth with MPDs remains lacking, despite their potential to yield critical insights into how risk and protective environmental influences on neurobehavioral development.

Building on these body of evidence, we developed an integrative transdiagnostic framework to jointly characterize environmental exposures, behavioral phenotypes, and normative brain structural deviations in youth with MPDs. Specifically, we included 1,755 youths aged 10-24 years, comprising individuals with MPDs and healthy controls (HCs), and examined 19 environmental exposures across four domains (5 positive and 12 negative social environments, and one positive and one negative natural environments), gray matter volume (GMV) centiles for 34 normative brain structural measures, and 40 behavioral measures across four domains (16 emotional symptoms, 8 non-emotional symptoms, 10 cognitive measures, and 6 personality and maladaptive belief measures). Guided by three hypotheses, our analytical workflow (Fig. 1) proceeded as follows. First, we tested whether behavioral, neuroanatomical, and environmental profiles exhibit transdiagnostic gradients alongside diagnosis-specific deviations. Second, using sparse canonical correlation analysis (sCCA)^40,41^, we tested whether latent brain-behavior modes capture shared variance across disorders and extend beyond traditional diagnostic boundaries. Third, we examined environmental influences by testing whether behavior mediates associations between environmental exposures and brain structure, and that environmental contexts moderate brain-behavior coupling, with these pathways differing across diagnostic groups.

**Fig. 1.**
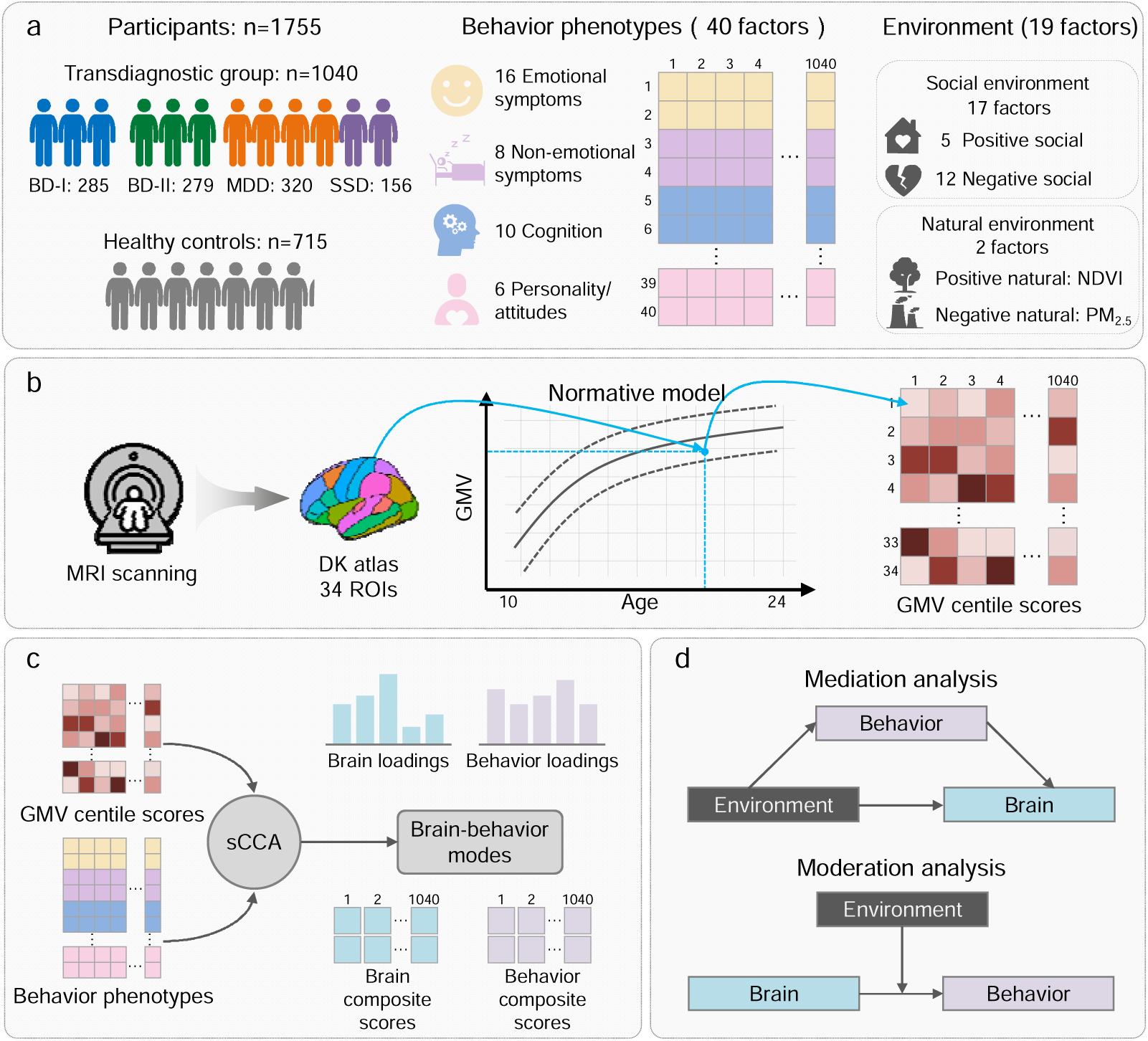
Schematic overview of the analytical framework. **a** Participant enrollment and assessment. Behavioral traits and social environmental factors were assessed using self-report and clinician-administered formats. Natural environmental factors were obtained from satellite remote sensing data. **b** Gray matter volume (GMV) centile scores quantification. Individual GMV centile scores were quantified using Lifespan Brain Chart Consortium (LBCC) framework^21^. **c** Multivariate correlation analysis. Sparse canonical correlation analysis (sCCA) was applied to identify optimal linear combinations of brain and behavioral features that maximized their correlation. **d** Mediation of the environment-brain relationship by behavioral phenotype, and moderation of the brain-behavior relationship by environment.

## Results

### Experimental design

This study integrated behavioral, neuroanatomical, and environmental data from 1,755 youth aged 10-24 years, including 285 with BD-Type I (BD-I), 279 with BD-Type II (BD-II), 320 with MDD, 156 with SSD, and 715 HCs. Demographic and clinical characteristics are summarized in Table S1. The multidimensional data comprised behavioral phenotyping, including emotional and non-emotional symptoms, cognitive function, and personality and maladaptive attitudes/beliefs; GMV centile scores quantified using the Lifespan Brain Chart Consortium (LBCC) framework^21^; and comprehensive environmental measures, encompassing 12 negative and 5 positive social factors (parental behaviors, childhood trauma, social support) as well as two natural exposures (PM_2.5_ and the Normalized Difference Vegetation Index, *NDVI*). We aimed to: (1) characterize transdiagnostic profiles and diagnosis-specificity across behavioral, neuroanatomical, and environmental dimensions; (2) identify multivariate brain-behavior relationships that transcend conventional diagnostic categories; and (3) test whether environmental factors relate to, mediate, or moderate these brain-behavior associations.

### Transdiagnostic severity gradients across behavioral phenotypes

Analysis of variance (ANOVA) revealed significant between-group differences across all four behavioral phenotypes (all *P*_FDR_ < 0.001; Fig. 2a, Table S2). For each of the four behavioral dimensions, the principal component analysis (PCA) was performed to extract its first principal component (PC1). The resulting PC1 scores for each dimension were then compared across the diagnostic groups (Fig. 2a).

**Fig. 2.**
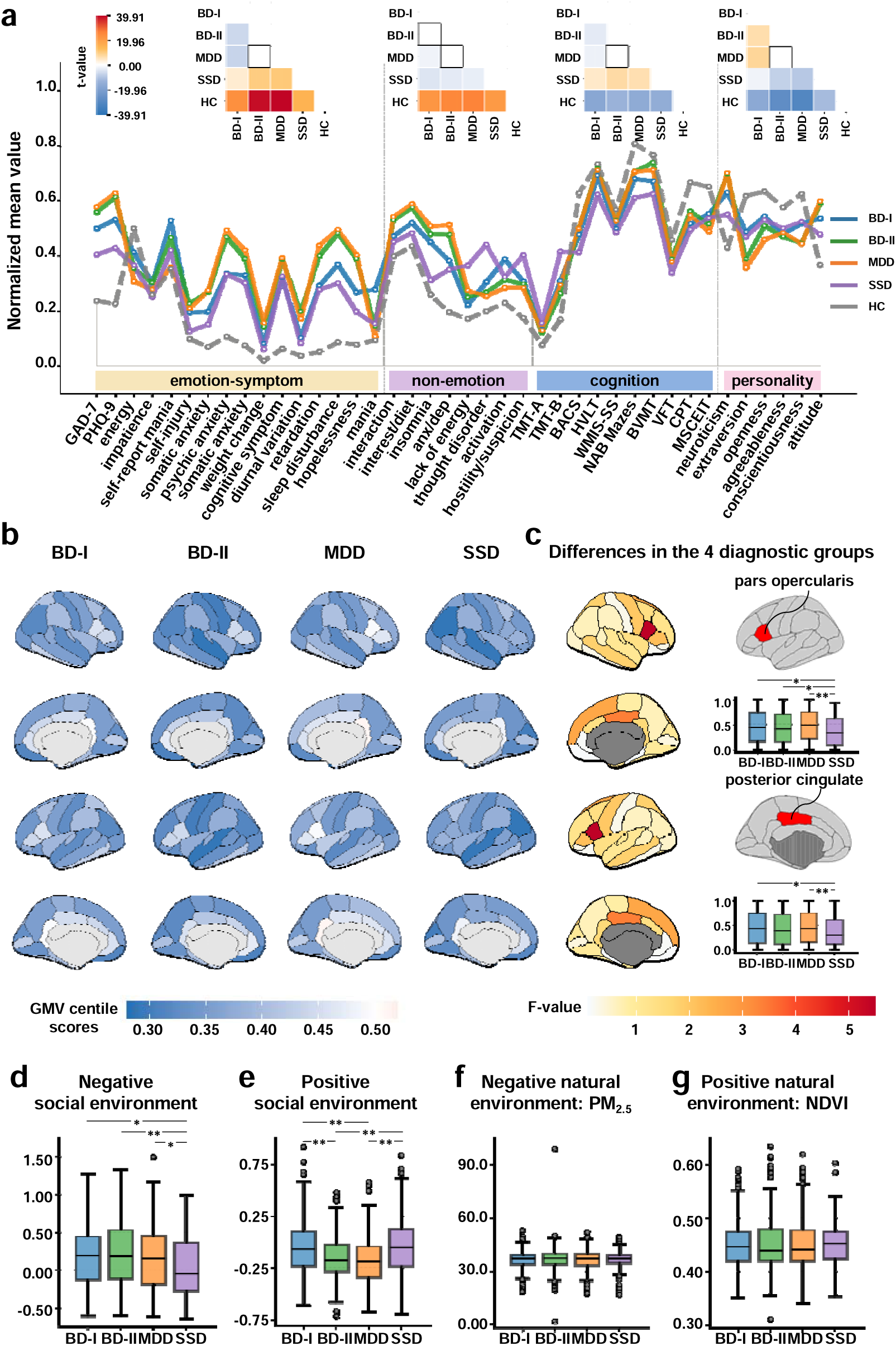
Comparisons of behavioral, neuroanatomical, and environmental profiles across diagnostic groups. **a** Severity gradient of behavioral phenotypes. Lower panel: Normalized mean values of the original behavioral phenotypes per diagnostic group. Upper panel: Pairwise comparison of behavioral PC1 scores across groups (white-circled squares indicate non-significant comparisons). PC1 (first principal component) was derived separately for each behavioral dimension within each group. **b** Regional GMV centile scores from the normative model^42^. Scores range from 0 to 1, with 0.5 indicating the reference sample median (50th centile). **c** Regional between-group differences in GMV centile scores. F-statistics from a one-way ANOVA identified two regions with significant differences: the pars opercularis (SSD < BD-I = BD-II = MDD) and the posterior cingulate (SSD < BD-I/MDD). **d-g** Group differences in social and natural environmental factors. **(d)** Negative social, **(e)** positive social, **(f)** negative natural, and **(g)** positive natural. All patient groups were demographically matched (age and gender) to controls using propensity score matching. Note: Asterisks denote statistical significance: \**P*_FDR_ < 0.05, \*\**P*_FDR_ < 0.001. The HCL-32 (energy, impatience), HAMA (somatic, psychic anxiety), HAMD (somatic anxiety, weight change, cognitive deficit, diurnal variation, retardation, sleep disturbance, and hopelessness), SHAPS (interactions, interest/diet), BPRS (anxiety/depression, lack of energy, thought disorder, activation, hostility/suspicion), and NEO (neuroticism, extraversion, agreeableness, openness, conscientiousness).

In emotion-related symptoms, all patient groups scored significantly higher than HCs on nearly all measures (all *P*_FDR_ < 0.001, Table S2). A severity gradient was observed for PC1 across patient groups, with BD-II and MDD patients scoring highest (all *P*_FDR_ < 0.001), followed by BD-I patients, and SSD patients scoring lowest (Fig. 2a).

For non-emotional symptoms, patients also scored significantly higher than HCs (all *P*_FDR_ < 0.001, Table S2). SSD showed the highest PC1 score (all *P*_FDR_ < 0.001), differing significantly from BD-I, BD-II, and MDD (Fig. 2a).

Cognitive performance also varied across groups. All patient groups demonstrated impairments relative to HCs (all *P*_FDR_ < 0.001, Table S2), with SSD exhibiting the poorest performance (all *P*_FDR_ < 0.001), followed by BD-I, and then BD-II and MDD (Fig. 2a).

For personality and attitudinal traits, patient groups exhibited elevated levels of Neuroticism and Dysfunctional Attitudes (with MDD and BD-II scoring highest, followed by BD-I and then SSD), alongside reduced scores on Extraversion, Openness, Agreeableness, and Conscientiousness (Fig. 2a, Table S2).

### Shared and disorder-specific deviations in GMV

To characterize transdiagnostic GMV profiles, we calculated centile scores (range: 0-1, where 0.5 represents the normative median (50th centile)) from the normative model for the four disorders. All patient groups showed widespread negative GMV deviations relative to the normative model, with highly similar spatial profiles across groups (Fig. 2b). ANOVA revealed two regions with significant between-group differences (Fig. 2c). Specifically, the pars opercularis showed significantly greater reductions in SSD than in BD-I, BD-II, and MDD (all *P*_FDR_ < 0.05, Table S3), whereas the posterior cingulate displayed stronger negative deviations in SSD compared with BD-I and MDD (all *P*_FDR_ < 0.05, Table S3). These findings indicate a shared pattern of cortical GMV loss across diagnoses, alongside disorder-specific abnormalities in select regions.

### Transdiagnostic gradients in social but not natural environments

Environmental profiles differed significantly across diagnostic groups for social environments, but not for natural environments. PCA was applied respectively to the positive and negative social environments domains, and PC1 scores were compared across diagnostic groups (Fig. 2d-e). For the negative social environment, SSD scored significantly lower than BD-I, and MDD (all *P*_FDR_ < 0.05), and even lower than BD-II (*P*_FDR_ < 0.001, Fig. 2d). For the positive social environment, SSD and BD-I exhibited significantly higher scores than BD-II and MDD (all *P*_FDR_ < 0.001; Fig. 2e). The natural environments showed no difference across different diagnostic conditions (Fig. 2f-g). Patient groups exhibited distinct environmental profiles compared to HCs across both social and natural environments (Fig. S1, Table S4-5).

### Multivariate brain-behavior relationships identified by sCCA

To examine multivariate associations between brain structure and behavior, sCCA was applied to 34 GMV centile measures and 40 behavioral variables. Based on the inflection point of the explained covariance curve, the first 8 of 34 modes were retained (Fig. 3a-b). Permutation testing identified two significant canonical modes: Mode 1 (*r* = 0.19, *P*_FDR_ = 0.008) was characterized by Cognitive Functions, while Mode 2 (*r* = 0.16, *P* = 0.032) was characterized by Emotional Symptoms. Examination of the highest-loading features showed that Mode 1 was dominated by the BACS score, and Mode 2 by the NSSI score (Fig. 3c-d).

**Fig. 3.**
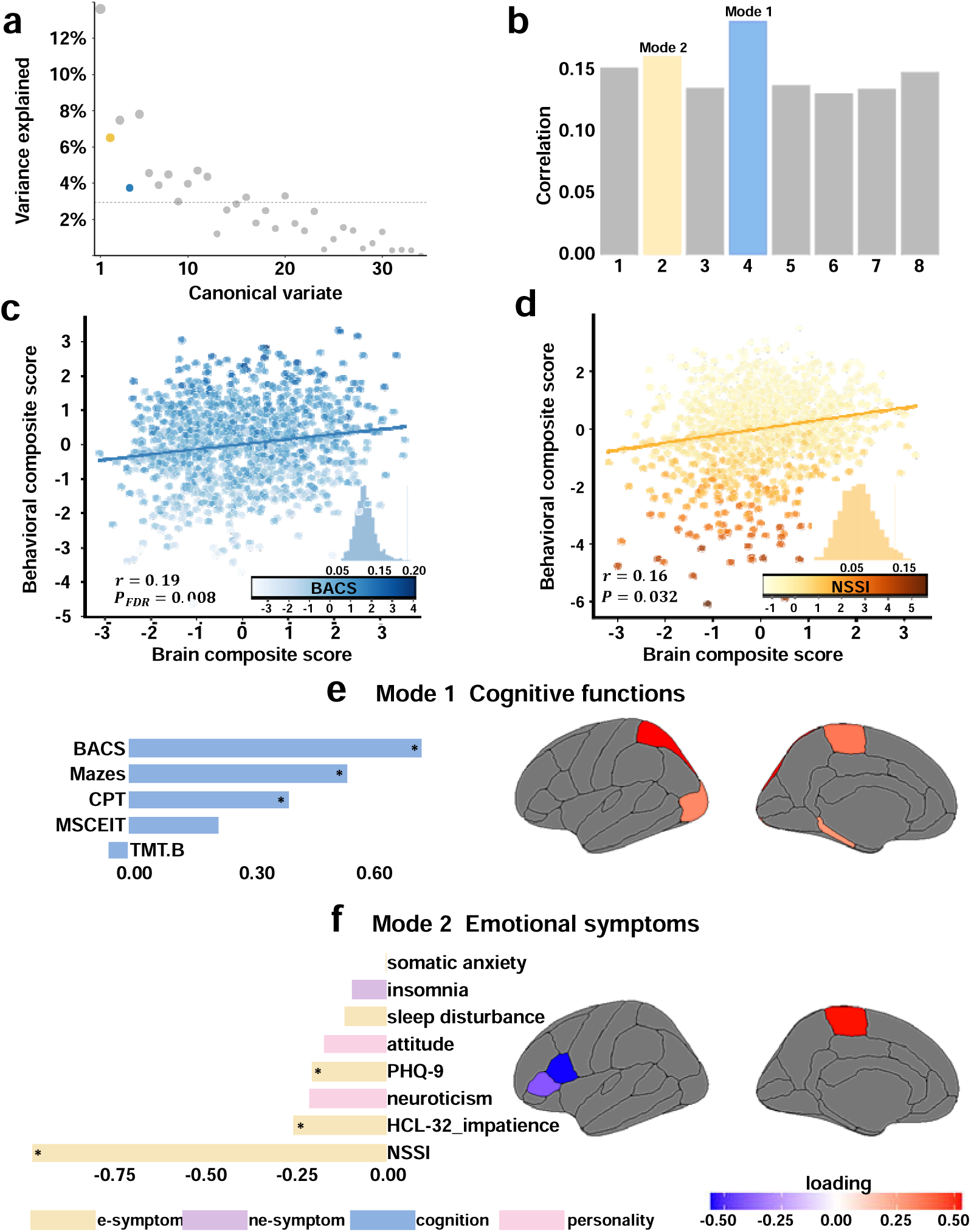
Multivariate brain-behavior relationships identified by sCCA. **a** Explained covariance of the first eight canonical modes. The dashed horizontal line indicates the mean explained covariance. **b** Statistical significance of each mode assessed by permutation testing with FDR correction. Mode 1: *P*_FDR_ < 0.01; Mode 2: P < 0.05 (uncorrected). Significant modes are highlighted in bright shades. **c-d** Scatter plots of brain composite scores (x-axis) against behavior composite scores (y-axis) for Mode 1 **(c)** and Mode 2 (**d**). Each point represents one participant; color intensity corresponds to the severity of the dominant clinical behavioral feature in each mode. **e** Behavioral and brain loadings for Mode 1 (“Cognitive Functions”), linking lower GMV centile scores in the lateral occipital cortex, superior parietal lobule, paracentral lobule, and parahippocampal gyrus to poorer performance in processing speed (BACS), reasoning/problem-solving (Mazes), and attention/vigilance (CPT). **f** Behavioral and brain loadings for Mode 2 (“Emotional Symptoms”), revealing that lower GMV centile score in the paracentral lobule were associated with higher severity of depression (PHQ-9), hypomania (HCL-32_impatience), and non-suicidal self-injury (NSSI); conversely, those in the pars opercularis and pars triangularis were associated with milder manifestations. Insets: Asterisks indicate features that have significant loadings based on bootstrap resampling (95% CI ≠0).

Mode 1 (“Cognitive Functions”) was associated with performance in several cognitive domains, including speed of processing, reasoning and problem solving, and attention/vigilance (Fig. 3e, Fig. S2). Bootstrap resampling identified three behavioral features with stable loadings (95% CI≠0): BACS (loading = 0.71), Mazes (loading = 0.53), CPT (loading = 0.39). This mode was reliably supported by four brain regions, primarily located in the lateral occipital cortex (loading = 0.38), superior parietal lobule (loading = 0.58), paracentral lobule (loading = 0.41), and parahippocampal gyrus (loading = 0.34). Together, these results suggest that lower GMV centile scores of the four brain regions are associated with poorer performance in specific cognitive functions: processing speed (assessed by BACS), reasoning and problem solving (reflected in Mazes), and attention/vigilance (measured by CPT).

Mode 2 (“Emotional Symptoms”) was characterized by features related to depression, hypomania, and self-injury (Fig. 3f, Fig. S3). Three behavioral features demonstrated stable loadings: NSSI (loading = -0.91), HCL-32_Impatience (loading = -0.24), PHQ-9 (loading = -0.19). This mode involved three stable brain regions, mainly located in paracentral lobule (loading = 0.52), pars opercularis (loading = -0.55), and pars triangularis (loading = -0.41). These findings indicated distinct, region-specific associations: lower GMV centile scores in the paracentral lobule with more severe symptoms, and in the pars opercularis and pars triangularis with milder forms of depression, hypomania, and self-injury.

Composite scores for both brain and behavior in both Mode 1 (Fig. S4a) and Mode 2 (Fig. S4b) exhibited clear divergence across diagnostic groups.

To evaluate the stability and generalizability of the identified multivariate brain-behavior relationships, we applied sCCA to a split-sample dataset comprising discovery (n = 697) and validation (n = 343) samples. Both samples showed comparable distributions in age, sex, and diagnosis (Fig. S5). The multivariate brain-behavior modes identified by sCCA in the discovery sample are presented in Fig. S6, with the resampling distributions for behavioral and brain features of Modes 1-3 shown in Figs. S6-9, respectively. Corresponding results from the validation sample are shown in Fig. S10 (modes) and Figs. S11-12 (resampling distributions for Modes 1 and 2). The canonical patterns derived from the discovery and validation samples were highly consistent with those obtained from the full sample, confirming the robustness of the multivariate brain-behavior relationships.

### Associations between environmental factors and behavioral phenotypes

Patterns of association between social environments and behavioral phenotypes differed across diagnostic groups (Fig. 4a). In BD-I, both positive and negative social environments were significant associated with a broad spectrum of emotional and non-emotional symptoms, personality and attitude traits. Specifically, positive social environments correlated negatively with emotional and non-emotional symptoms, and positively with adaptive personality and attitude traits, but showed no significant association with cognitive functions. Negative social environments exhibited the opposite pattern. This suggests that supportive social contexts may alleviate symptoms and promote adaptive personality traits and positive attitudes, whereas adverse conditions exacerbate them.

**Fig. 4.**
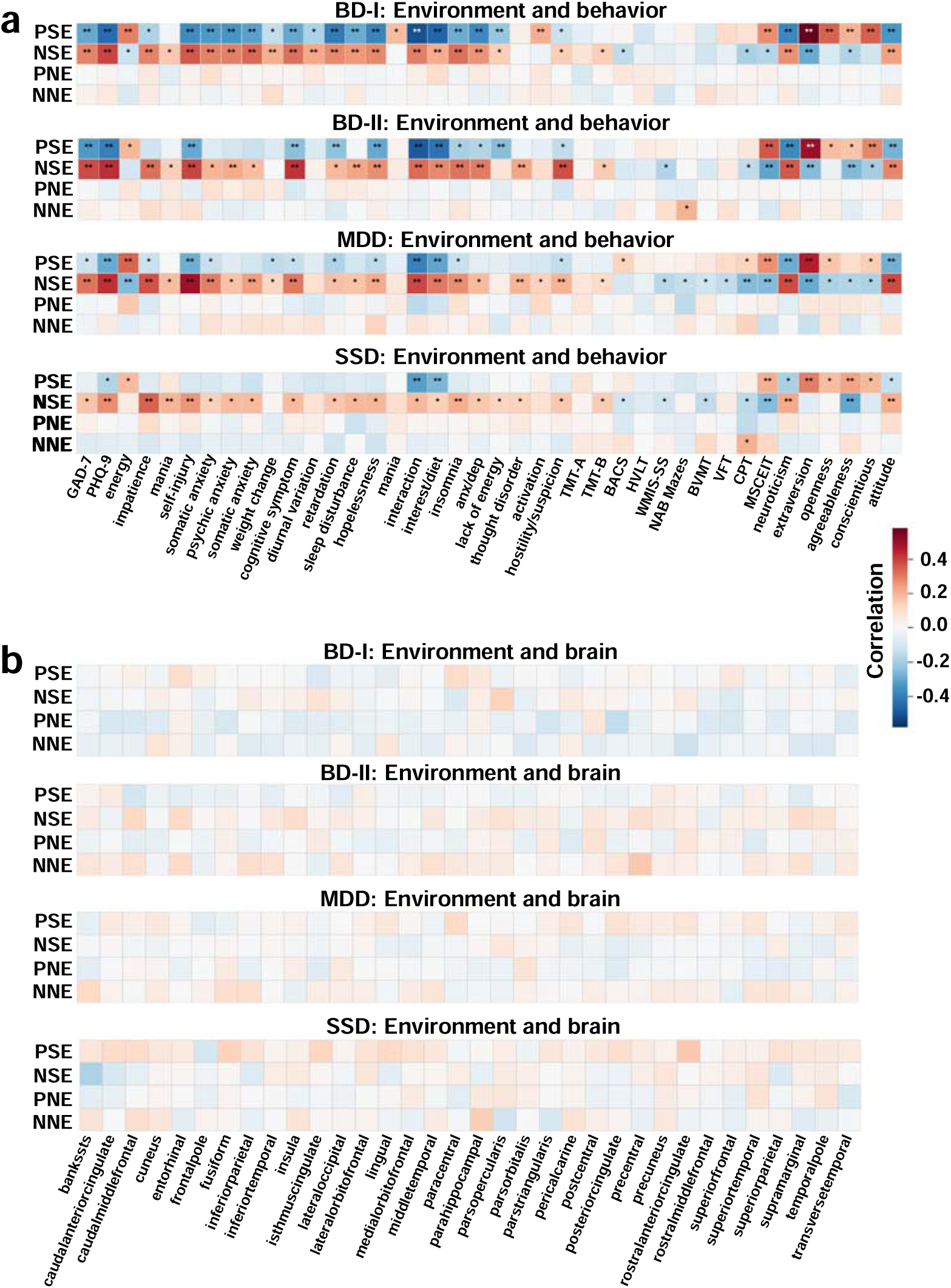
Correlations of environmental factors with behavioral and neuroanatomical measures. **a** Spearman’s correlations matrix between four environmental factors (PSE, positive social; NSE, negative social; PNE, positive natural; NNE, negative natural) and behavioral features. **b** Spearman’s correlations matrix between the same environmental factors and GMV centile scores of 34 brain regions. Note: Heatmaps depict the direction and strength of Spearman’s correlation coefficients (red, positive; blue, negative). Asterisks indicate significance correlations after FDR correction (*P < 0.05, **P < 0.001).

In BD-II and MDD, negative social environments showed broader and stronger associations with emotional and non-emotional symptoms and personality/attitude traits than positive environments. This indicates that social adversity exerts a widespread influence on behavioral dysregulation in both disorders. Notably, the association between negative social environments and cognitive impairment was more extensive in MDD than in BD-II, suggesting a heightened cognitive sensitivity to social adversity in MDD.

In SSD, negative social environments were significantly correlated with a wide range of behavioral measures, whereas positive social environments showed minimal correlations. This pattern indicates an attenuated behavioral responsiveness to positive social cues, alongside a heightened sensitivity to negative social conditions in SSD.

Associations between natural environments and behavioral phenotypes were largely absent across the four disorders (Fig. 4a). Only two significant correlations were observed: in BD-II, between PM_2.5_ and NAB Mazes, and in SSD, between PM_2.5_ and CPT.

### Associations between environmental factors and neuroanatomy

We found no significant associations between any of the four environmental factors and GMV centile scores in any of the 34 brain regions across the four diagnostic groups (Fig. 4b).

### Mediation and Moderation Effects

We performed causal mediation analysis (CMA) to test whether behavior mediates the effect of environments on the brain (environment→behavior→brain). Across 32 tested pathways spanning four environmental domains (positive and negative social and natural environments), two sCCA-derived modes, and four diagnostic groups (MDD, BD-I, BD-II, and SSD), eight mediation effects remained significant after False Discovery Rate (FDR) correction (95% CI excluding zero; Fig. 5a, b).

**Fig. 5.**
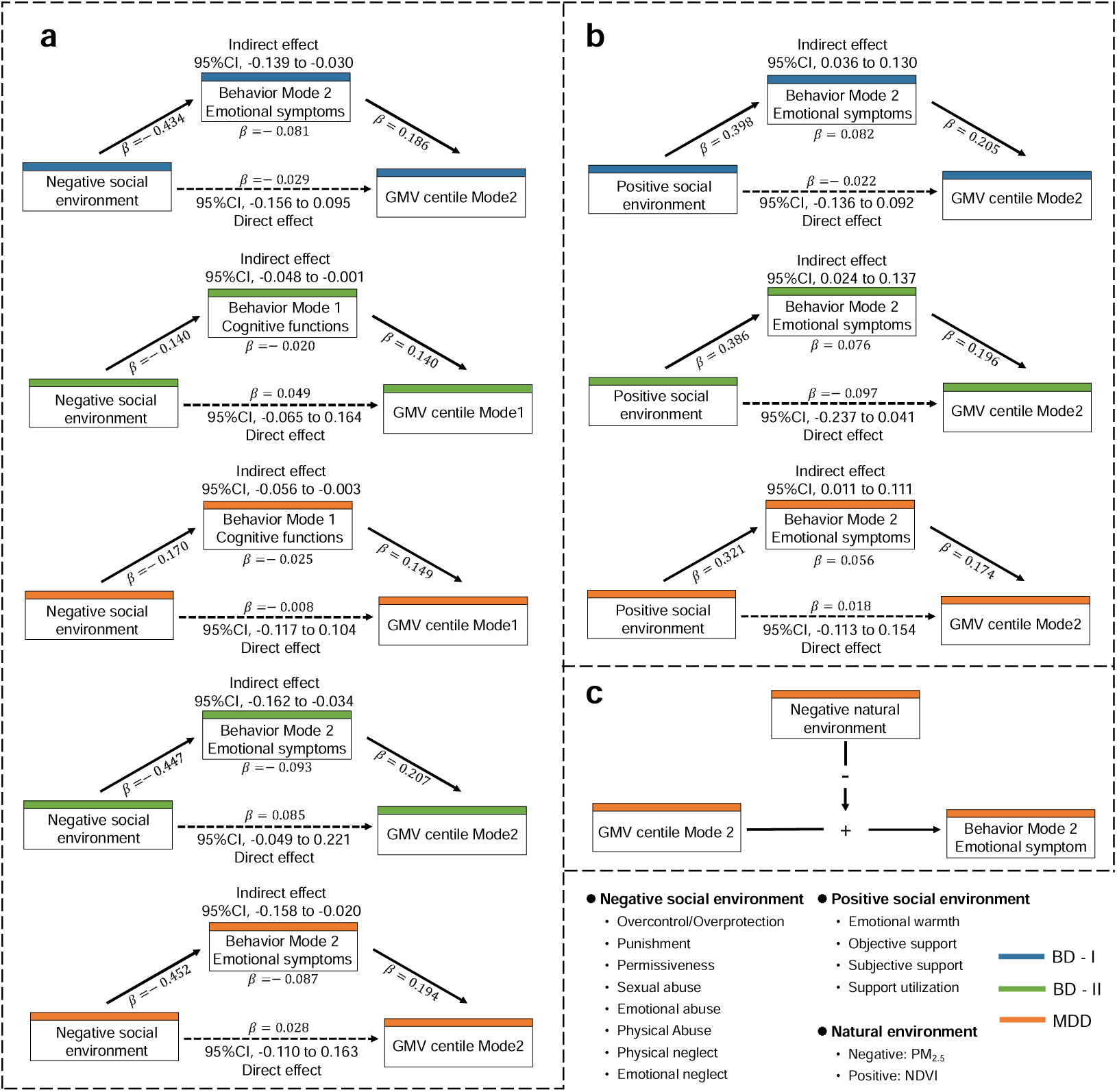
Mediation and Moderation Effects. **a-b** Schematics illustrating the mediation by behavioral composite scores (Modes 1 and 2) of the association between social environments and brain composite scores. Panel **(a)** shows the effect for the negative social environment, and panel **(b)** for the positive social environment, across participants with BD-I, BD-II, and MDD. **c** Moderation by the negative natural environments of the association between GMV centile scores and Mode 2 behavior in MDD.

In the environment→behavior→brain pathway, more adverse negative social environments were associated with lower behavior composite scores in Mode 1 (Cognitive Functions), which subsequently predicted lower brain composite scores in BD-II and MDD (Fig. 5a). For Mode 2 (Emotional symptoms), a more adverse negative social environments were associated with lower behavioral composite scores, which in turn predicted lower brain composite scores in BD-I, BD-II, and MDD (Fig. 5a). Similarly, an insufficient positive social environments were associated with lower behavioral composite scores in Mode 2, subsequently predicting lower brain composite scores in BD-I, BD-II, and MDD (Fig. 5b). No significant mediation was observed in participants with SSD in either pathway.

We further examined moderation effects and found that a more adverse negative natural environment significantly attenuated the positive association between brain and behavioral composite scores for Mode 2 (Emotional Symptoms) in MDD (Fig. 5c).

## Discussion

Investigating transdiagnostic gradients and interrelationships across behavioral, neuroanatomical, and environmental dimensions in youth with MPDs, using a combination of normative brain modeling, multivariate machine learning, and causal inference approaches, we present three major findings: (1) Behavioral, neuroanatomical, and environmental profiles show both transdiagnostic continuity and diagnosis-specific gradients across MPDs. (2) sCCA reveals multiple multivariate patterns of GMV centiles associated with transdiagnostic behavioral dimensions of psychopathology. (3) We identified diagnosis- and dimension-specific pathways in which behavioral dimensions statistically mediated associations between environmental exposures and brain structure in BD and MDD, whereas exposure to the negative natural environments (PM_2.5_) moderated brain-behavior relationships in MDD.

Consistent with our hypothesis of transdiagnostic continuity and diagnosis-specific gradients of neuroanatomical domain, our investigation revealed widespread negative deviation of GMV compared to normative brain charts across all four patient groups, as well as diagnosis-specific deviation in two regions: the pars opercularis and the posterior cingulate (Fig. 2b). The widespread negative deviations in GMV observed across diagnostic categories suggest that MPDs may share a common pattern of neural impairment. Previous studies have reported structural alterations in the pars opercularis and posterior cingulate cortex in schizophrenia^43^, MDD^44^ and BD^45^. Importantly, the two regions are key hubs of the action-mode network (AMN), previously labelled the cingulo-opercular network, which is critical for coordination of decision-making, goal maintenance, alertness, preparation and initiation of motor responses, as well as feedback processing (motor, physiological and body states, pain)^46^. In the present study, SSD showed the largest GMV deviations in these regions, suggesting a greater alteration of neural systems supporting action-oriented behavior. Consistent with this interpretation, previous studies have shown that disruption of the AMN is associated with apathy, abulia, and reduced self-initiated activity^47,48^, symptoms that are especially pronounced in SSD.

The substantial heterogeneity observed within each diagnostic category suggests that current symptom-based diagnostic criteria may not adequately “carve nature at its joints”^49^. This limitation has motivated the development of the RDoC framework, which extends discrete diagnostic characterization by integrating dimensional and categorical perspectives, thereby better capturing overlapping clinical features across disorders and facilitating the identification of core brain-behavior relationships that transcend traditional diagnostic boundaries^15^. Within this framework, our behavioral analyses revealed transdiagnostic severity gradients across multiple domains, including emotional and non-emotional symptoms, cognition, and personality traits, with diagnostic groups distributed along shared continuous dimensions rather than forming discrete clusters (Fig. 2a). Accordingly, an increasing body of research has focused on linking brain circuits^41,50^ or morphology^51^ to behavioral phenotypes across traditional diagnostic boundaries.

In the present study, we applied sCCA to identify two robust and clinically interpretable brain-behavior modes linking dimensional psychopathology to GMV centile scores across diagnostic categories. The first mode was characterized by a distributed posterior cortex encompassing the lateral occipital cortex, superior parietal lobule, paracentral lobule, and parahippocampal gyrus, in which lower GMV centile scores were associated with impairments in core cognitive domains, including processing speed, reasoning and problem solving, and attention/vigilance (Fig. 3e). These regions collectively support object processing, memory, and somato-motor integration^52–55^, suggesting that this mode reflects a transdiagnostic cognitive vulnerability associated with posterior cortical structural variation during neurodevelopment.

By contrast, the second mode delineated an emotional dimension involving both prefrontal regulatory regions, particularly the pars opercularis and pars triangularis, as well as the paracentral lobule, a sensorimotor region increasingly implicated in affective and pain-related processes. The pars opercularis and pars triangularis, core components of the inferior frontal gyrus, are critically involved in top-down regulation of affective responses^56^. Within this mode, reduced GMV centiles in these prefrontal regions were associated with relatively milder depressive, hypomanic, and self-injurious symptoms, whereas structural reductions in the paracentral lobule were linked to more severe emotional manifestations (Fig. 3f), supporting a severity-dependent neuroanatomical pattern of emotional dysregulation. This dissociation suggests that preserved structural integrity of prefrontal regulatory regions may partially buffer against symptom escalation, while greater involvement of sensorimotor-associated regions, particularly the paracentral lobule, which plays a role in the emotional modulation of pain perception^57^, may reflect more pervasive affective disturbance and help contextualize the observed association with self-injurious symptoms.

A notable feature shared by both brain-behavior modes is the paracentral lobule, formed in part by the medial extension of the primary motor cortex (M1), a key component of the somato-cognitive action network (SCAN). Through SCAN, this region shows strong connectivity with the AMN^58^, situating it at the intersection of sensorimotor processing and action control. The paracentral lobule supports cognitive functions related to attentional allocation and the translation of cognitive goals into organized actions, consistent with its association with cognitive impairments observed in the first mode. Beyond cognition, the paracentral lobule has also been implicated in emotional regulation^59^. Its involvement across both modes suggests that structural variation in this region may link cognitive and emotional dimensions of psychopathology via shared SCAN-AMN circuitry.

Importantly, the pars opercularis contributed prominently to the second brain-behavior mode and was also among the regions showing diagnosis-specific GMV deviations across all four patient groups, with the most pronounced reductions observed in SSD. This convergence suggests that the pars opercularis may represent a key node linking transdiagnostic emotional dimensions with disorder-specific neurodevelopmental variation.

A growing body of evidence indicates that early-life exposure to social and natural environments contributes to mental health risk^60–65^. We derived four environmental domains: positive social, negative social, positive natural, and negative natural by applying PCA to 17 social (5 positive, 12 negative) factors and 2 natural indicators (NDVI and PM_2.5_). This integrative approach extends prior studies that examined isolated environmental exposures^62,66^, enabling a more comprehensive characterization of environmental burden across diagnostic groups. Using this framework, we identified diagnosis-specific patterns of social environmental exposure and their differential associations with behavioral dimensions (Fig. 4a). In BD-I, both positive and negative social environments broadly correlated with symptoms, personality and maladaptive attitudes/beliefs, suggesting generalized social-emotional reactivity^67^. In BD-II and MDD, associations centered predominantly on adverse social environments, which are in agreement with the stress model^68,69^. Conversely, SSD showed stronger associations with negative social environments and comparatively weaker associations with positive social factors, potentially reflecting impairments in social cognition and emotion recognition^70^.

The absence of significant direct associations between environmental factors and regional GMV suggests that early-life environmental influences on brain structure during youth may not operate through simple linear pathways (Fig. 4b). Instead, these findings point toward indirect mechanisms whereby environmental exposures are preferentially expressed at the behavioral level, with downstream associations with neuroanatomical variation^38^. Accordingly, environmental influences on brain structure may be better captured through coordinated brain-behavior dimensions rather than direct structure-environment associations.

Our mediation analyses revealed diagnosis- and dimension-specific environmental contributions to brain-behavior relationships across the two sCCA-derived modes, formally delineating these indirect pathways. Dimension-specific analyses demonstrated distinct mediational pathways (Fig. 5a-b). Greater exposure to negative social environments and reduced exposure to positive social environments were associated with poorer cognitive performance in Mode 1, which in turn mediated lower GMV centile scores in posterior cortical regions. Conversely, the same environmental risk profiles were associated with milder emotional symptoms in Mode 2, which subsequently mediated reduced GMV centiles in emotion-related regions. Collectively, these findings indicate that social environments influence brain structure through differentiated behavioral pathways rather than a uniform mechanism. Consistent with this interpretation, previous studies have shown that psychiatric problems mediate the association between family conflict and reduced cortical areas in children^38^, and that the brain continuously rewires itself in response to learning and life experiences^49^. Extending this framework, our results demonstrate that social environments may shape neuroanatomical variation indirectly by modulating specific behavioral dimensions in youths with MPDs.

Diagnosis-specific analyses further underscored heterogeneity in these pathways. In BD-I, BD-II, and MDD, social environmental adversity exerted its neuroanatomical associations predominantly through behavioral mediation, consistently with models of psychosocial experiences becoming biologically embedded^71^. In contrast, no significant behavioral mediation was observed in SSD, suggesting that neuroanatomical alterations in the schizophrenia spectrum may be less dependent on acquired social environments and may be primarily shaped by intrinsic neurodevelopmental and genetic vulnerability, consistent with the high heritability of schizophrenia (approximately 80%)^72^.

Beyond mediation, we identified a diagnosis-specific moderating role of the natural environment. In MDD, higher exposure to air pollution (PM_2.5_) attenuated the positive association between brain structure and the emotional symptoms dimension (Mode 2; Fig. 5c). Mechanistically, animal studies suggest that air pollutants can access the central nervous system through barrier dysfunction, triggering neuroinflammatory and autoimmune responses^73^, and emerging evidence links such pollution-related neuroimmune alterations to disrupted brain-behavior relationships involved in emotional regulation^74^. In contrast, no significant moderating effects of either the natural or social environment on brain-behavior relationships were observed in BD or SSD. Moreover, social environments did not show a moderating effect in any diagnostic group, consistent with the notion that social exposures may exert cumulative neurodevelopmental influences by shaping behavioral phenotypes^38^. Together, these findings suggest that brain-behavior relationships in MDD are particularly sensitive to adverse natural environmental perturbations, consistent with the relatively lower heritability of MDD (approximately 40%)^75^ and its greater environmental susceptibility.

Several limitations should be considered when interpreting our findings. Although our environmental measures were comprehensive, certain aspects of exposure may remain unaccounted for, particularly for natural environments where finer-grained assessments could reveal subtler effects. Moreover, residential-based estimates, even with high-resolution satellite data, may not fully capture individual mobility or local microenvironmental variation. In addition, due to model constraints, cortical measures were averaged across hemispheres. Given that MPDs have been associated with atypical hemispheric asymmetry^76,77^; our findings should therefore be interpreted in light of this potential limitation, as they are based on a symmetrical (unihemispheric average) representation of the cerebral hemispheres. Finally, the cross-sectional nature of the current dataset precludes direct causal inferences regarding the associations of environmental exposures with brain and behavioral phenotypes.

In summary, this study establishes a framework that integrates brain structural MRI with comprehensive behavioral and environmental exposure measures in a well-characterized large-sample clinical cohort, and demonstrates both transdiagnostic and disorder-specific neurobehavioral gradients in youth with MPDs. We identified diagnosis-specific neuroanatomy deviations in key hubs of the AMN: pars opercularis and posterior cingulate cortex, and also delineating two transdiagnostic brain-behavior modes (cognitive and emotional) that converge on the paracentral lobule, a core region within the SCAN. Moreover, we further dissected distinct environmental pathways: social environment adversity influenced brain morphometry alterations through behavior in BD and MDD, whereas exposure to negative natural environments (PM_2.5_) moderated brain-behavior relationships specifically in MDD. Collectively, these findings advance understanding of transdiagnostic neurobehavioral gradients in MPDs and highlight how social and natural environmental factors are differentially embedded in brain-behavior associations. Building upon this multidimensional framework, a pivotal next step will be to bridge the gap between neurobehavioral dimensions across diagnostic boundaries and precision psychiatry.

### Methods

#### Participants

A total of 1,755 participants aged 10-24 years were enrolled in this study, including 285 individuals with BD-I, 279 with BD-II, 320 with MDD, 156 with SSD, and 715 HCs. Patient recruitment was conducted in the inpatient psychiatry unit of Renmin Hospital of Wuhan University between March 2023 and March 2025. This unit serves as the largest psychiatric department within a general hospital in Central China. All diagnoses were established by trained psychiatrists using the Structured Clinical Interview for DSM-5, Patient Edition (SCID-I/P). Exclusion criteria for patients included neurological disorders, severe physical illness, substance abuse, severe verbal/visual impairments, braces, or claustrophobia. The enrolled participants originated from nearly all regions of China, with a majority concentrated in Hubei Province (Fig. S13).

HCs were recruited via advertisements in schools and local communities across Hubei Province and neighboring provinces. Exclusion criteria for HCs comprised a personal or family history of psychiatric disorders, significant physical or neurological illness, and severe verbal or visual impairments. The absence of a personal psychiatric history was confirmed for all HCs using the Structured Clinical Interview for DSM-5, Non-Patient Edition (SCID-I/NP).

Written informed consent was obtained from all participants and/or their legal guardians. The study was approved by the Institutional Review Board of Wuhan University (WHU-LFMD-IRB2023021) and was registered with the Chinese Clinical Trials Registry (ChiCTR2500106094).

#### Assessment of emotional and non-emotional symptoms

Emotional symptoms included: anxiety, measured by the Hamilton Anxiety Scale (HAMA)^78^ and the Generalized Anxiety Disorder-7 (GAD-7)^79^; depression, evaluated with the Hamilton Depression Rating Scale-24 (HAMD-24)^80^ and Patient Health Questionnaire-9 (PHQ-9)^81^; mania, assessed using the Young Mania Rating Scale (YMRS)^82^ and Mood Disorder Questionnaire (MDQ)^83^; hypomania, measured by the Hypomania Checklist (HCL-32)^84^; and non-suicidal self-injury, evaluated via the Chinese version of the Adolescent Non-suicidal Self-injury Questionnaire (NSSI) (Table S6).

Non-emotional symptoms comprised: anhedonia, measured by the Snaith-Hamilton Pleasure Scale (SHAPS)^85^; insomnia, assessed using the Insomnia Severity Index (ISI)^86^; and general psychiatric symptoms, evaluated with the Brief Psychiatric Rating Scale (BPRS)^87^.

For scales with established factor structures, factor scores were used; otherwise, total scores were applied. The HCL-32 consists of two factors: Energy and Impatience. HAMA comprises two factors: Somatic and Psychic Anxiety. HAMD includes seven factors: Anxiety/Somatization, Weight Change, Cognitive Deficit, Diurnal Variation, Retardation, Sleep Disturbance, and Hopelessness. SHAPS contains two factors: Interactions/Experiences and Interest/Diet. BPRS is divided into five factors: Anxiety/Depression, Lack of Energy, Thought Disorder, Activation, and Hostility/Suspicion. Comprehensive details are provided in Tables S7 and S8.

#### Assessment of cognitive functions

Cognitive functions was assessed using the MATRICS Consensus Cognitive Battery-Chinese version (MCCB) ^88,89^, covering seven domains: speed of processing, as evaluated by the Trail Making Test parts A and B (TMT-A/B), Verbal Fluency Test (VFT), and BACS-Symbol Coding; visual learning and memory, using the Brief Visuospatial Memory Test (BVMT); working memory, assessed by the Wechsler Memory Scale Spatial Span (WMS-SS); vigilance and attention, measured with the Continuous Performance Test (CPT); verbal learning and memory, via the Hopkins Verbal Learning Test (HVLT); problem-solving and reasoning, using the NAB Mazes; and social cognition, with the Mayer-Salovey-Caruso Emotional Intelligence Test (MSCEIT). Detailed information are presented in Table S9. All assessments were administered by trained examiners, and inter-rater reliability exceeded 0.85 for all cognitive measures.

#### Assessment of personality and maladaptive attitudes/beliefs

Personality traits were assessed using the NEO Personality Inventory^90^, which measures five factors: Neuroticism, Extraversion, Agreeableness, Openness, and Conscientiousness (Table S10). Maladaptive attitudes/beliefs were evaluated using the Dysfunctional Attitudes Scale^91^ (Table S10).

#### Assessment of positive and negative social environment

Maternal and paternal parenting styles were assessed using the Memories of Parental Rearing Behaviour (MPRB) questionnaire^92^ (Table S11). The maternal parenting style comprises four factors: Emotional Warmth, Overcontrol/Overprotection, Punishment, and Permissiveness. The paternal parenting style includes five factors: Emotional Warmth, Overcontrol, Punishment, Overprotection, and Permissiveness. Emotional Warmth represents positive social environment, with higher scores indicating a more supportive environment. The remaining factors represent negative social environment, where higher scores reflect a less favorable parenting environment.

Social Support: Participants’ social support was evaluated using the Social Support Rating Scale (SSRS)^93^, which includes three factors: Objective Support, Subjective Support, and Utilization of Social Support (Table S12). All three factors represent positive social environment, where higher scores indicating a more supportive social environment.

Childhood trauma was evaluated using the Childhood Trauma Questionnaire (CTQ)^94^, which consists of five factors: Sexual Abuse, Emotional Abuse, Physical Abuse, Physical Neglect, and Emotional Neglect (Table S13). These factors represent negative social environment, with higher scores indicating a less favorable childhood environment.

The PC1 derived from the PCA of five positive and twelve negative social environmental factors was used to represent the respective social environments. Age and sex were included as covariates in the analyses.

#### Assessment of positive and negative natural environment

This study assessed exposure to the natural environment using remote sensing data with extensive spatial coverage from 2018 to 2025. For each participant, exposure was defined as the five-year period preceding enrollment. Average daily PM_2.5_ concentrations and monthly NDVI values during this window, derived from satellite observations, served as indicators of air pollution (negative exposure) and vegetation cover (positive exposure), respectively. All datasets were resampled to a 100-meter resolution to support street-scale analysis. Auxiliary datasets, including meteorological variables, were incorporated to enhance the accuracy of PM_2.5_ mapping. Detailed methods for PM_2.5_ concentration, NDVI, and exposure variable extraction are described in the Supplementary Methods.

#### MRI acquisition and preprocessing

T1-weighted (T1W) sagittal images were obtained with a 3D T1W Bravo sequence on a GE SIGNA 3T scanner in the Radiology Department of Renmin Hospital, Wuhan University. The images underwent surface-based preprocessing with the FreeSurfer software package (https://surfer.nmr.mgh.harvard.edu/). GMV was automatically derived for each cortical region. Subsequently, GMV values from the 68 cortical structures were residualized against total intracranial volume (TIV), sex, and age using a linear regression model. Detailed protocols for T1W image acquisition and preprocessing are provided in the Supplementary Methods.

#### Brain centile score extraction

Individualized GMV centile scores were derived for 34 bilateral regions from the Desikan-Killiany atlas. These regions were formed by merging homologous structures from the original 68 cortical structures, and the scores were computed using the Lifespan Brain Chart Consortium (LBCC) framework^21^. Preprocessed regional GMV measures were projected onto the sex-stratified neurodevelopmental trajectories via Generalized Additive Models for Location, Scale and Shape (GAMLSS)^95^. For each participant and region, we computed a centile score (range 0-1) through maximum likelihood estimation against the reference curves. These scores quantify an individual’s GMV position relative to the normative population distribution, where a score of 0.5 corresponds to the population 50th centile. Scanner and site effects were inherently adjusted for within the covariance structure of the reference model.

To examine systematic departures from normative expectations, we performed one-sample t-tests on the centile scores for each diagnostic group. This analysis produced regional t-statistics that quantified the magnitude and direction of group-level deviations from the healthy normative model.

We quantified regional cortical GMV using the Desikan-Killiany atlas and converted values into centile scores based on a normative model. For each diagnostic group, regional mean centile scores were computed and projected onto the cortical surface for visualization. Group differences in regional GMV centile scores were assessed using ANOVA across 34 bilateral cortical regions, followed by pairwise Welch’s t-tests with FDR correction.

#### Comparisons of behavioral, environmental, and neuroanatomical measures

We first conducted univariate analyses to characterize group differences in behavioral, environmental, and neuroanatomical measures across diagnostic groups and HCs. Behavioral and neuroanatomical variables were compared using ANOVA followed by post-hoc tests. For environmental variables, we first employed propensity score matching to create demographically matched HCs for each diagnostic group based on age and sex. Group comparisons were then conducted using two-sample t-tests between each patient group and its individually matched HCs. All comparisons were corrected for multiple testing using the FDR (Benjamini-Hochberg procedure).

#### Sparse canonical correlation analysis

We employed sCCA, an unsupervised learning technique, to model the multivariate associations between regional GMV centile scores and behavioral phenotypes. The analysis was implemented using the R PMA package following a previous study^41^, with elastic net regularization applied separately to the brain and behavior data. Prior to the analysis, the influence of covariates (age and sex) were regressed out from the behavioral data. The sCCA derived linear combinations of brain and behavioral features that maximized their correlation, with optimal tuning parameters selected through 10-fold cross-validation. Statistical significance was assessed through permutation testing (1,000 iterations) with FDR correction for multiple comparisons.

The stability of the resulting modes was evaluated via bootstrap resampling (1,000 iterations). Features with confidence intervals not crossing zero were considered stable.

#### Correlation analyses

To assess pairwise associations, we performed Spearman’s rank correlation analyses between each environmental factor and regional GMV centile scores or behavioral features within each diagnostic group. The FDR method was applied to correct for multiple comparisons.

#### Mediation and moderation analyses

To investigate whether behavior mediates the effect of environment on the brain, we conducted CMA for each significant sCCA mode. The models included environmental factors as independent variables, behavior composite scores (from sCCA) as mediators, and brain composite scores (from sCCA) as outcomes. Analyses were performed using the R mediation package^96^ with a quasi-Bayesian Monte Carlo approach (5,000 simulations). A significant mediation effect was defined as a 95% confidence interval for the average causal mediation effect (ACME) that excluded zero.

We also performed moderation analysis to test whether environmental factors influence the relationship between GMV composite scores and behavioral scores. For each significant brain-behavior mode, a linear regression model^97^ was fitted with the behavioral composite score as the outcome (Y). The model incorporated the GMV composite score (X), the environmental moderator (M), their interaction term (X×M), and age and sex as covariates. The model is expressed as:

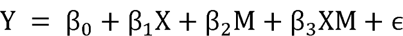

Where β0 is the intercept, β1 represents the effect of the brain feature (X) on behavior (Y), β2 indicates the main effect of the environmental moderator (M), β3 quantifies the interaction effect between X and M (i.e., the moderation effect), and Є denotes the error term. All variables were mean-centered prior to analysis. The significance of interaction terms was assessed with FDR correction applied within each diagnostic group.

## Reporting summary

Further information on research design is available in the Nature Portfolio Reporting Summary linked to this article.

## Data availability

Requests for access to the data should be directed to the corresponding author.

## Code availability

Structural MRI data were preprocessed using FreeSurfer v6.0, which is publicly available at https://surfer.nmr.mgh.harvard.edu/. Information regarding members of the Lifespan Brain Chart Consortium is available at https://docs.google.com/spreadsheets/d/1D8YNDcnhwlv2WcUDhreq3fwrkfpfGiFp0OGFVO5d-es/edit?usp=sharing. Codes for this research is available at https://github.com/Yeyi-mindlab/YMPDs/tree/main, where the sCCA algorithm code was based on Author Xia C H, Ma Z, Ciric R, et al., (2018).

## Acknowledgements

The study was supported by the National Key R&D Program of China (2024YFC3308400), National Natural Science Foundation of China (62576254, 62201356, 32361143787), Fundamental Research Funds for the Central Universities (2042025YXA006), Major Project of Science and Technology Innovation of Hubei Province (2024BCA003), Hubei Provincial Natural Science Foundation (2024AFB751), Key Project of Hubei Provincial Health Commission (WJ2023Z003), the Shenzhen-Hong Kong Institute of Brain Science-Shenzhen Fundamental Research Institutions (2023SHIBS0003), Medicine Plus Program of Shenzhen University (2024YG021), Undergraduate Training Programs for Innovation of Wuhan University (202510486145), and Natural Science Foundation of Xiaogan City (XGKJ2024010061, XGKJ2024010062, XGKJ2024010063 and XGKJ2025010002).

## Author contributions

Authors Xiaofen Zong, Yi Ye, Jinxin He, Kaitong Ma, Mang Ye, Tao Yao, Siwei Li, He Li, Ge Song, Yinshan Wang, Mengyao Feng, Qi Wen, Jie Yao, Li Dong, Yuanyuan Zhang, Xia Sun, Bing Xiang Yang, Maolin Hu, Xinian Zuo, Xujun Duan and Li Zhang were involved in participant recruitment, and clinical data collection and analysis. Xiaofen Zong, Yi Ye, Siwei Li, Ge Song, Yinshan Wang, Xinian Zuo, Xujun Duan and Li Zhang performed the analysis. Xiaofen Zong, Xujun Duan and Li Zhang designed the study and supervised all aspects of the research. Xiaofen Zong, Yi Ye and Maolin Hu wrote the first draft of the manuscript. Xiaofen Zong, Yi Ye, Jinxin He, Xujun Duan and Li Zhang revised the manuscript. All authors contributed to and approved the final version of the paper.

