## Supplementary for "Transdiagnostic Neurobehavioral Gradients and Environmental Interactions in Youth with Major Psychiatric Disorders"

Catalogues

Supplementary Results ... 3
Table S1. Demographic and clinical characteristics of patients and healthy controls ... 3
Table S2. Comparisons of behavior phenotypes (emotional and non-emotional symptoms, cognition, and personality/attitude) among groups ... 4
Table S3. Comparisons of neuroanatomical profiles among groups ... 10
Table S4. Comparisons of social psychological environment between patient groups and healthy controls ... 13
Table S5. Comparisons of natural environment between patient groups and healthy controls ... 14
Fig. S1 Group differences in social and natural environment factors ... 15
Fig. S2 Resampling distributions for behavioral and brain features for Mode 1 in the full sample ... 16
Fig. S3 Resampling distributions for behavioral and brain features for Mode 2 in the full sample ... 16
Fig. S4 Transdiagnostic group differences in canonical brain-behavior modes ... 17
Fig. S5 Demographic characteristics of the discovery and validation samples ... 17
Fig. S6 Multivariate brain-behavior relationships identified by sCCA in the discovery sample. ... 18
Fig. S7 Resampling distributions of behavioral and brain features for Mode 1 in the discovery dataset ... 19
Fig. S8 Resampling distributions of behavioral and brain features in Mode 2 in the discovery dataset ... 20
Fig. S9 Resampling distributions of behavioral and brain features in Mode 3 in the discovery dataset. Features in bold have 95% confidence intervals that do not cross zero, indicating their stable contributions to the mode. ... 20
Fig. S10 Multivariate brain-behavior relationships identified by sCCA in the validation sample. a Explained covariance of the first three canonical modes ... 21
Fig. S11 Resampling distributions of behavioral and brain features in Mode 1 in the validation dataset ... 22
Fig. S12 Resampling distribution of behavioral and brain features in Mode 2 in the validation dataset ... 23
Fig. S13 Geographical distribution of sampling locations ... 23
Supplementary Methods ... 24
Table S6. Description of Adolescent Non-suicidal Self-injury Questionnaire ... 24
Table S7. Description of emotion-related clinical symptoms ... 25
Table S8. Description of non-emotion-related clinical symptoms ... 27
Table S9. Description of MATRICS Consensus Cognitive Battery ... 28
Table S10. Description of personality and maladaptive attitudes/beliefs ... 29
Table S11. Description of social psychological environment: parental parenting style ... 31
Table S12. Description of Social Support Rating Scale ... 34
Table S13. Description of Childhood Trauma Questionnaire ... 36

### Supplementary Results

**Table S1**. Demographic and clinical characteristics of patients and healthy controls

| Clinical data | Disease | BD-I | BD-II | MDD | SSD | HCs | $t/\chi^2$ | $P^a$ | $F/\chi^2$ | $P^b$ |
|---|---|---|---|---|---|---|---|---|---|---|
| N | 1040 | 285 | 279 | 320 | 156 | 715 | - | - | - | - |
| Age, years | 16.62±3.005 | 17.07±2.925 | 16.43±2.845 | 16.17±3.021 | 17.10±3.234 | 16.41±2.310 | 1.608 | 0.092 | 6.338 | ＜0.001 |
| Female/Male | 682/358 | 182/103 | 210/69 | 218/102 | 72/84 | 455/260 | 0.699 | 0.403 | 39.263 | ＜0.001 |
| Education[c], years | 10.09±2.779 | 10.43±2.726 | 10.05±2.733 | 9.76±2.821 | 10.19±2.810 | 10.25±2.179 | -1.344 | 0.179 | 3.158 | 0.013 |
| First episode, yes/no, n | 874/166 | 215/70 | 238/41 | 280/40 | 141/15 | - | - | - | - | - |
| Disease Duration[d], months | 24.94 | 27.84 | 28.14 | 23.04 | 17.79 | - | - | - | - | - |
| Drug naïve, n | 565 | 128 | 135 | 195 | 107 | - | - | - | - | - |
| Drug free[e], n | 475 | 157 | 144 | 125 | 49 | - | - | - | - | - |
| Prior psychotropic drug use within the drug-free group | | | | | | | | | | |
| Antipsychotics, no. | 254 | 94 | 75 | 48 | 37 | - | - | - | - | - |
| Antidepressants, no. | 233 | 65 | 77 | 77 | 14 | - | - | - | - | - |
| Mood stabilizers, no. | 181 | 80 | 60 | 32 | 9 | - | - | - | - | - |
| Hypnotics, no. | 65 | 17 | 19 | 24 | 5 | - | - | - | - | - |

Note: Disease: Disease group; BD-I: Bipolar disorder type I; BD-II: Bipolar disorder type II; MDD, Major Depressive Disorder; SSD: Schizophrenia spectrum and other psychotic disorders; HCs: Healthy controls. *t*, Independent Samples t-test; $\chi^2$, Chi-square test; *F,* one-way ANOVA; *N*, the sample size of a certain group.

[a]*P* value for comparisons between Disease group and HCs.

[b]*P* value for comparisons among BD-I, BD-II, MDD, SSD and HCs.

c: Data were missing for educational years in two patients.

d: Data were missing for disease duration in six patients.

e: Drug free status in patients was defined as no exposure to oral psychotropics for at least 6 weeks or depot psychotropics for 6 months.

**Table S2.** Comparisons of behavior phenotypes (emotional and non-emotional symptoms, cognition, and personality/attitude) among groups

| Behaviors | BD-I, n=285 | BD-II, n=279 | MDD, n=320 | SSD, n=156 | HCs, n=715 | F | $P_{FDR}$ | Post hoc | | |
|---|---|---|---|---|---|---|---|---|---|---|
| | | | | | | | | comparison | 4 diseases groups vs HCs | 4 diseases groups |
| GAD-7_Total score | 10.33±6.05 | 11.96±5.33 | 12.33±5.55 | 7.87±6.15 | 4.29±3.94 | 208.46 | <0.001 | MDD=BD-II>BD-I>SSD>HC | Ps<0.001 | BD-I vs BD-II: P<0.05; BD-II vs MDD: P>0.05; Remaining comparisons: Ps<0.001 |
| PHQ-9_Total score | 14.55±7.57 | 17.38±6.47 | 17.72±6.09 | 11.01±7.27 | 5.26±4.38 | 371.36 | <0.001 | MDD=BD-II>BD-I>SSD>HC | Ps<0.001 | BD-II vs MDD: P>0.05; Remaining comparisons: Ps<0.001 |
| HCL-32_Energy | 11.04±6.07 | 7.99±6.10 | 6.00±5.62 | 9.33±6.06 | 14.13±6.37 | 130.87 | <0.001 | HC>BD-I>SSD=BD-II>MDD | Ps<0.001 | BD-I vs SSD: P<0.05; BD-II vs SSD: P>0.05; Remaining comparisons: Ps<0.001 |
| HCL-32_Impatience | 5.58±2.64 | 5.49±2.56 | 4.88±2.50 | 4.46±2.73 | 4.47±5.17 | 6.35 | <0.001 | BD-I=BD-II>MDD=HC=SSD | HC vs MDD HC vs SSD: Ps>0.05; Remaining comparisons: Ps<0.001 | BD-I vs SSD BD-II vs SSD: Ps<0.001; BD-I vs BD-II: P>0.05; Remaining comparisons: Ps<0.05 |
| MDQ_Total score | 6.70±3.26 | 5.82±3.07 | 4.32±2.51 | 5.21±3.47 | 4.27±2.61 | 45.18 | <0.001 | BD-I>BD-II=SSD>MDD=HC | HC vs SSD: P<0.05; HC vs MDD: P>0.05; Remaining comparisons : | BD-I vs BD-II MDD vs SSD: Ps<0.05; SSD vs BD-II: P>0.05; Remaining comparisons: Ps<0.001 |

| | | | | | | | | | | |
|---|---|---|---|---|---|---|---|---|---|---|
| | | | | | | | | | Ps<0.001 | |
| NSSI_ Total score | 16. 08± 21. 72 | 22.25 ±23.5 7 | 19.7 7±23 .23 | 5.24 ±13. 60 | 2.93±6. 63 | 103 .02 | <0. 001 | BD-II=MDD=BD-I>SSD >HC | Ps<0.001 | BD-I vs BD-II: P<0.05; BD-II vs MDD BD-I vs MDD: Ps>0.05; Remaining comparisons: Ps<0.001 |
| HAMA_Somatic Anxiety | 4.6 8±5 .60 | 6.77± 4.89 | 6.85 ±5.4 4 | 3.23 ±4.7 5 | 1.21±2. 35 | 138 .16 | <0. 001 | MDD= BD-II>BD-I>SSD >HC | Ps<0.001 | BD-I vs SSD: P<0.05; MDD vs BD-II: P>0.05; Remaining comparisons: Ps<0.001 |
| HAMA_Psychological Anxiety | 8.7 0±6 .01 | 12.45 ± 4.62 | 13.0 3±4. 82 | 8.41 ±5.9 7 | 2.44± 3.21 | 420 .15 | <0. 001 | MDD= BD-II>BD-I=SSD >HC | Ps<0.001 | BD-II vs MDD BD-I vs SSD: Ps>0.05; Remaining comparisons: Ps<0.001 |
| HAMD-24_Anxiety/ Somatization | 3.9 8±2 .80 | 4.88± 2.30 | 5.27 ±2.6 3 | 3.51 ±2.6 5 | 0.57±0. 99 | 406 .85 | <0. 001 | MDD> BD-II>BD-I=SSD >HC | Ps<0.001 | MDD vs BD-II: P<0.05; BD-I vs SSD: P>0.05; Remaining comparisons: Ps<0.001 |
| HAMD-24_Weight Change | 0.1 4±0 .45 | 0.27± 0.57 | 0.29 ±0.6 1 | 0.09 ±0.3 5 | 0.01±0. 13 | 34. 47 | <0. 001 | MDD= BD-II>BD-I=SSD >HC | HC vs SSD: P<0.05; Remaining comparisons : Ps<0.001 | BD-I vs BD-II: P<0.05; BD-I vs SSD BD-II vs MDD: Ps>0.05; Remaining comparisons: Ps<0.001 |
| HAMD-24_Cognitive Deficit | 5.1 1±3 .50 | 6.46± 3.31 | 6.53 ±3.1 7 | 5.09 ±3.6 9 | 0.43±0. 87 | 470 .23 | <0. 001 | MDD= BD-II>BD-I=SSD >HC | Ps<0.001 | SSDvs BD-II: P<0.05; BD-I vs SSD BD-II vs MDD: Ps>0.05; Remaining comparisons: |

| | | | | | | | | | | |
|---|---|---|---|---|---|---|---|---|---|---|
| | | | | | | | | | | Ps<0.001 |
| HAMD-24_Diurnal Variation | 0.16±0.46 | 0.37±0.63 | 0.32±0.60 | 0.11±0.40 | 0.03±0.17 | 42.57 | <0.001 | BD-II=MDD>BD-I=SSD>HC | HC vs SSD: P<0.05; Remaining comparisons: Ps<0.001 | BD-I vs SSD BD-II vs MDD: Ps>0.05; Remaining comparisons: Ps<0.001 |
| HAMD-24_Retardation | 3.21±2.49 | 4.60±1.88 | 5.03±1.83 | 2.95±2.47 | 0.31±0.69 | 584.98 | <0.001 | MDD>BD-II>BD-I=SSD>HC | Ps<0.001 | BD-II vs MDD: P<0.05; BD-I vs SSD: P>0.05; Remaining comparisons: Ps<0.001 |
| HAMD-24_Sleep Disturbance | 2.20±1.94 | 2.97±1.79 | 3.03±1.80 | 1.70±1.90 | 0.37±0.81 | 251.16 | <0.001 | MDD=BD-II>BD-I>SSSD>HC | Ps<0.001 | BD-I vs SSD: P<0.05; BD-II vs MDD: P>0.05; Remaining comparisons: Ps<0.001 |
| HAMD-24_Hopelessness | 2.60±2.45 | 4.05±2.18 | 4.17±2.00 | 1.72±2.03 | 0.49±0.86 | 342.36 | <0.001 | MDD=BD-II>BD-I>SSD>HC | Ps<0.001 | BD-II vs MDD: P>0.05; Remaining comparisons: Ps<0.001 |
| YMRS_Total score | 10.20±9.20 | 3.58±4.53 | 1.78±3.16 | 4.40±5.57 | 1.27±3.62 | 154.67 | <0.001 | BD-I>SSD=BD-II>MDD>HC | HC vs MDD: P<0.05; Remaining comparisons: Ps<0.001 | BD-II vs SSD: P>0.05; Remaining comparisons: Ps<0.001 |
| SHAPS_Interactions/Experiences | 12.24±3.76 | 13.86±3.45 | 14.32±3.49 | 11.86±3.66 | 10.50±5.02 | 64.80 | <0.001 | MDD=BD-II>BD-I=SSD>HC | Ps<0.001 | BD-I vs SSD BD-II vs MDD: Ps>0.05; Remaining comparisons: Ps<0.001 |
| SHAPS_Interest/Diet | 16.83±5.28 | 18.77±4.36 | 19.37±4.47 | 15.84±5.08 | 14.31±6.47 | 67.56 | <0.001 | MDD=BD-II>BD-I>SSD>HC | HC vs SSD: P<0.05; Remaining comparisons: | SSD vs BD-I: P<0.05; BD-II vs MDD: P>0.05; Remaining |

| | | | | | | | | | | |
|---|---|---|---|---|---|---|---|---|---|---|
| | | | | | | | | | Ps<0.001 | comparisons: Ps<0.001 |
| ISI_Total score | 12.32±7.45 | 13.17±6.81 | 13.97±6.73 | 8.11±6.43 | 6.59±5.19 | 114.83 | <0.001 | MDD=BD-II=BD-I>SSD>HC | HC vs SSD: P<0.05; Remaining comparisons: Ps<0.001 | MDD vs BD-I: P<0.05; BD-I vs BD-II BD-II vs MDD: Ps>0.05; Remaining comparisons: Ps<0.001 |
| BPRS_Anxiety/Depression | 9.06±4.04 | 11.38±3.52 | 12.15±3.63 | 8.13±4.20 | 4.48±2.46 | 401.10 | <0.001 | MDD>BD-II>BD-I=SSD>HC | Ps<0.001 | MDD vs BD-II: P<0.05; BD-I vs SSD: P>0.05; Remaining comparisons: Ps<0.001 |
| BPRS_Lack of Energy | 5.17±1.76 | 5.69±2.07 | 6.24±2.57 | 8.57±3.71 | 3.90±2.05 | 157.95 | <0.001 | SSD>MDD>BD-II>BD-I>HC | Ps<0.001 | MDD vs BD-II: P<0.05; Remaining comparisons: Ps<0.001 |
| BPRS_Thought Disorder | 6.22±2.69 | 5.33±1.77 | 5.03±2.02 | 9.05±3.67 | 3.97±2.06 | 164.85 | <0.001 | SSD>BD-I>BD-II=MDD>HC | Ps<0.001 | BD-II vs MDD: P>0.05; Remaining comparisons: Ps<0.001 |
| BPRS_Activation | 5.11±1.98 | 4.07±1.37 | 3.66±1.04 | 4.29±1.64 | 2.99±1.56 | 102.12 | <0.001 | BD-I>SSD=BD-II>MDD>HC | Ps<0.001 | MDD vs SSD: P<0.05; BD-II vs SSD: P>0.05; Remaining comparisons: Ps<0.001 |
| BPRS_Hostility/Suspicion | 5.23±2.34 | 5.02±1.86 | 4.76±1.89 | 6.84±2.56 | 2.97±1.56 | 183.17 | <0.001 | SSD>BD-I=BD-II=MDD>HC | Ps<0.001 | MDD vs BD-I: P<0.05; BD-II vs MDD BD-II vs BD-I: P>0.05; Remaining comparisons: Ps<0.001 |

| | | | | | | | | | | |
|---|---|---|---|---|---|---|---|---|---|---|
| MCCB_TMT-A | 48.533±20.286 | 45.75±18.55 | 47.43±19.60 | 54.05±35.03 | 32.36±13.70 | 79.15 | <0.001 | SSD=BD-I=MDD=BD-II>HC | Ps<0.001 | BD-I vs SSD MDD vs BD-I MDD vs BD-II: Ps>0.05; Remaining comparisons: Ps<0.05 |
| MCCB_TMT-B | 109.28±59.17 | 97.40± 49.20 | 106.40±57.66 | 139.23±81.13 | 69.90±40.83 | 78.18 | <0.001 | SSD>BD-I=MDD>BD-II>HC | Ps<0.001 | BD-I vs BD-II MDD vs BD-II: Ps<0.05; BD-I vs MDD: P>0.05; Remaining comparisons: Ps<0.001 |
| MCCB_BACS-SC | 52.64±11.21 | 54.79±10.76 | 54.66±10.93 | 46.56±13.00 | 66.87±9.82 | 199.93 | <0.001 | HC>BD-II=MDD>BD-I>SSD | Ps<0.001 | MDD vs BD-I BD-I vs BD-II: Ps<0.05; BD-II vs MDD: P>0.05; Remaining comparisons: Ps<0.001 |
| MCCB_HVLT | 24.89±5.11 | 25.78±4.67 | 25.61±4.49 | 22.436±6.04 | 26.36±4.09 | 24.97 | <0.001 | HC=BD-II=MDD=BD-I>SSD | HC vs MDD: P<0.05; HC vs BD-II: P>0.05; Remaining comparisons: Ps<0.001 | BD-II vs BD-I: P<0.05; MDD vs BD-I MDD vs BD-II: Ps>0.05; Remaining comparisons: Ps<0.001 |
| MCCB_WMS-SS | 14.56±3.04 | 15.22±3.18 | 15.18±3.05 | 14.32±3.29 | 16.29±2.78 | 29.60 | <0.001 | HC>BD-II>MDD=BD-I=SSD | Ps<0.001 | SSD vs BD-II: P<0.001; BD-I vs SSD BD-II vs MDD: Ps>0.05; Remaining comparisons: Ps<0.05 |
| MCCB_NA | 18.19± | 18.67±5.37 | 18.78±5.41 | 16.66±6.51 | 21.60±4.34 | 53.06 | <0.001 | HC>MDD=BD- | Ps<0.001 | SSD vs MDD SSD vs BD-II: Ps<0.001; |

| | | | | | | | | | | |
|---|---|---|---|---|---|---|---|---|---|---|
| B Mazes | 5.68 | | | | | | | II=BD-I>SSD | | BD-I vs SSD: P<0.05; Remaining comparisons: Ps>0.05 |
| MCCB_BVMT | 24.68±7.00 | 27.55±5.98 | 26.49±6.36 | 22.70±7.33 | 28.48±4.72 | 40.09 | <0.001 | HC>BD-II>MDD>BD-I>SSD | HC vs BD-II: P<0.05; Remaining comparisons: Ps<0.001 | MDD vs BD-I BD-I vs SSD MDD vs BD-II: Ps<0.05; Remaining comparisons: Ps<0.001 |
| MCCB_Fluency | 20.35±5.77 | 20.33±5.89 | 19.82±5.86 | 18.53±5.95 | 23.44±5.77 | 45.72 | <0.001 | HC>BD-I=BD-II=MDD>SSD | Ps<0.001 | BD-I vs SSD SSD vs MDD: P<0.05; SSD vs BD-II: P<0.001; Remaining comparisons: Ps>0.05 |
| MCCB_CPT | 1.84±0.87 | 2.02±0.85 | 1.94±0.86 | 1.76±0.92 | 2.60±0.69 | 92.300 | <0.001 | HC>BD-II=MDD>BD-I=SSD | Ps<0.001 | BD-I vs SSD BD-II vs MDD: Ps>0.05; SSD vs BD-II: P<0.001; Remaining comparisons: Ps<0.05 |
| MCCB_MSCEIT | 92.37±15.53 | 88.90±15.07 | 86.53±15.04 | 91.20±15.42 | 99.67±13.42 | 60.76 | <0.001 | HC>BD-I=SSD=BD-II=MDD | Ps<0.001 | BD-I vs MDD: P<0.001; MDD vs SSD BD-I vs BD-II: Ps<0.05; Remaining comparisons: Ps>0.05 |
| NEO_Neuroticism | 43.13±9.99 | 47.06±7.60 | 47.15±6.70 | 38.44±8.67 | 32.89±9.36 | 223.96 | <0.001 | MDD=BD-II>BD-I>SSD>HC | Ps<0.001 | BD-II vs MDD: P>0.05; Remaining comparisons: Ps<0.001 |
| NEO_Extraversion | 34.78± | 29.81±8.32 | 28.38±7.90 | 34.18±8.60 | 40.87±7.47 | 177.56 | <0.001 | HC>BD-I=SSD | Ps<0.001 | BD-II vs MDD: P<0.05; BD-I vs SSD: |

| | | | | | | | | | | |
|---|---|---|---|---|---|---|---|---|---|---|
| | 8.87 | | | | | | | >BD-II>MDD | | P>0.05; Remaining comparisons: Ps<0.001 |
| NEO_Openness | 40.45±7.67 | 38.60±7.80 | 36.33±7.50 | 40.00±6.79 | 44.21±6.88 | 77.51 | <0.001 | HC>BD-I=SSD=BD-II>MDD | Ps<0.001 | BD-I vs BD-II: P<0.05; BD-I vs SSD BD-II vs SSD: Ps>0.05; Remaining comparisons: Ps<0.001 |
| NEO_Agreeableness | 36.81±6.72 | 36.49±6.75 | 37.13±6.52 | 37.77±5.69 | 41.30±6.37 | 48.44 | <0.001 | HC>SSD=MDD=BD-I=BD-II | Ps<0.001 | BD-II vs SSD: P<0.05; Remaining comparisons: Ps>0.05 |
| NEO_Conscientiousness | 37.25±8.69 | 33.10±7.66 | 32.96± 7.36 | 37.81±7.66 | 42.34±7.99 | 114.83 | <0.001 | HC>SSD=BD-I>BD-II=MDD | Ps<0.001 | BD-I vs SSD BD-II vs MDD: Ps>0.05; Remaining comparisons: Ps<0.001 |
| DAS_Total score | 160.40 ±36.25 | 174.09±34.72 | 175.24±32.76 | 146.80 ± 29.08 | 125.50± 30.27 | 196.91 | <0.001 | MDD=BD-II>BD-I>SSD>HC | Ps<0.001 | BD-II vs MDD: P>0.05; Remaining comparisons: Ps<0.001 |

Note: HAMA, Hamilton Anxiety Scale; GAD-7, Generalized Anxiety Disorder-7; HAMD-24, Hamilton Depression Rating Scale-24; PHQ-9, Patient Health Questionnaire-9; YMRS, Young Mania Rating Scale; MDQ, Mood Disorder Questionnaire; HCL-32, Hypomania Checklist-32; NSSI, the Chinese version of the Adolescent Non-suicidal Self-injury Questionnaire; SHAPS, Snaith-Hamilton Pleasure Scale; ISI, Insomnia Severity Index; BPRS, Brief Psychiatric Rating Scale; NEO, NEO Personality Inventory; DAS, Dysfunctional Attitudes Scale.

**Table S3.** Comparisons of neuroanatomical profiles among disease groups

| **ROI-centile** | **BD-I (n=285)** | **BD-II (n=279)** | **MDD (n=320)** | **SSD (n=156)** | **F** | ***P*** | **Post hoc** | |
|---|---|---|---|---|---|---|---|---|
| | | | | | | | **comparisons** | **4 diagnostic groups** |
| pars opercularis | 0.46 ± 0.32 | 0.45 ± 0.31 | 0.49 ± 0.30 | 0.37 ± 0.29 | 5.48 | <0.05 | MDD=BD-II=BD-I>SSD | SSD vs BD-I SSD vs BD-II: Ps_adj<0.05; SSD vs MDD: P_adj<0.001; |

| | | | | | | | | |
|---|---|---|---|---|---|---|---|---|
| | | | | | | | | Remaining comparisons: Ps_adj>0.05 |
| posterior cingulate | 0.45 ± 0.32 | 0.42 ± 0.32 | 0.46 ± 0.32 | 0.37 ± 0.29 | 3. 37 | P_raw< 0.05; P_adj >0.05 | MDD = BD-I > SSD | SSD vs BD-I SSD vs MDD: Ps_adj<0.05; Remaining comparisons: Ps_adj>0.05 |
| bankssts | 0.37 ± 0.28 | 0.34 ± 0.26 | 0.37 ± 0.27 | 0.36 ± 0.29 | 0. 61 | >0.05 | BD-I=MDD=SSD=BD-II | Ps>0.05 |
| caudalanteriorcingulate | 0.44 ± 0.27 | 0.44 ± 0.28 | 0.46 ± 0.28 | 0.41 ± 0.27 | 1. 38 | >0.05 | MDD=BD-I=BD-II=SSD | Ps>0.05 |
| caudalmiddlefrontal | 0.40 ± 0.29 | 0.37 ± 0.30 | 0.41 ± 0.30 | 0.41 ± 0.29 | 1. 39 | >0.05 | MDD=SSD=BD-I=BD-II | Ps>0.05 |
| cuneus | 0.33 ± 0.29 | 0.32 ± 0.28 | 0.35 ± 0.30 | 0.30 ± 0.26 | 1. 02 | >0.05 | MDD=BD-I=BD-II=SSD | Ps>0.05 |
| entorhinal | 0.48 ± 0.26 | 0.45 ± 0.25 | 0.47 ± 0.27 | 0.49 ± 0.25 | 1. 08 | >0.05 | SSD=BD-I=MDD=BD-II | Ps>0.05 |
| frontalpole | 0.41 ± 0.30 | 0.36 ± 0.28 | 0.41 ± 0.31 | 0.37 ± 0.29 | 2. 06 | >0.05 | MDD=BD-I=SSD=BD-II | Ps>0.05 |
| fusiform | 0.41 ± 0.30 | 0.38 ± 0.27 | 0.41 ± 0.30 | 0.39 ± 0.29 | 0. 93 | >0.05 | BD-I=MDD=SSD=BD-II | Ps>0.05 |
| inferiorparietal | 0.32 ± 0.29 | 0.32 ± 0.28 | 0.33 ± 0.28 | 0.29 ± 0.27 | 0. 64 | >0.05 | MDD=BD-I=BD-II=SSD | Ps>0.05 |
| inferiortemporal | 0.43 ± 0.30 | 0.42 ± 0.28 | 0.43 ± 0.29 | 0.41 ± 0.28 | 0. 28 | >0.05 | MDD=BD-I=BD-II=SSD | Ps>0.05 |
| insula | 0.39 ± 0.31 | 0.38 ± 0.29 | 0.42 ± 0.31 | 0.38 ± 0.31 | 1. 36 | >0.05 | MDD=BD-I=SSD=BD-II | Ps>0.05 |

| | | | | | | | | |
|---|---|---|---|---|---|---|---|---|
| isthmuscingulate | 0.50 ± 0.34 | 0.45 ± 0.32 | 0.51 ± 0.32 | 0.48 ± 0.31 | 1.59 | >0.05 | MDD=BD-I=SSD=BD-II | Ps>0.05 |
| lateraloccipital | 0.34 ± 0.29 | 0.33 ± 0.28 | 0.36 ± 0.29 | 0.33 ± 0.27 | 0.52 | >0.05 | MDD=BD-I=BD-II=SSD | Ps>0.05 |
| lateralorbitofrontal | 0.34 ± 0.30 | 0.33 ± 0.28 | 0.34 ± 0.29 | 0.35 ± 0.30 | 0.19 | >0.05 | SSD=BD-I=MDD=BD-II | Ps>0.05 |
| lingual | 0.34 ± 0.26 | 0.34 ± 0.26 | 0.36 ± 0.28 | 0.32 ± 0.26 | 0.98 | >0.05 | MDD=BD-I=BD-II=SSD | Ps>0.05 |
| medialorbitofrontal | 0.31 ± 0.29 | 0.31 ± 0.28 | 0.32 ± 0.30 | 0.32 ± 0.30 | 0.12 | >0.05 | MDD=SSD=BD-I=BD-II | Ps>0.05 |
| middletemporal | 0.36 ± 0.31 | 0.33 ± 0.29 | 0.39 ± 0.31 | 0.35 ± 0.29 | 1.79 | >0.05 | MDD=BD-I=SSD=BD-II | Ps>0.05 |
| paracentral | 0.38 ± 0.29 | 0.35 ± 0.27 | 0.38 ± 0.29 | 0.37 ± 0.29 | 0.82 | >0.05 | BD-I=MDD=SSD=BD-II | Ps>0.05 |
| parahippocampal | 0.40 ± 0.26 | 0.41 ± 0.25 | 0.41 ± 0.27 | 0.40 ± 0.27 | 0.09 | >0.05 | MDD=BD-II=SSD=BD-I | Ps>0.05 |
| parsorbitalis | 0.38 ± 0.33 | 0.35 ± 0.33 | 0.37 ± 0.33 | 0.36 ± 0.32 | 0.29 | >0.05 | BD-I=MDD=SSD=BD-II | Ps>0.05 |
| parstriangularis | 0.44 ± 0.32 | 0.44 ± 0.31 | 0.46 ± 0.33 | 0.39 ± 0.30 | 2.16 | >0.05 | MDD=BD_I=BD_II=SSD | Ps>0.05 |
| pericalcarine | 0.43 ± 0.30 | 0.41 ± 0.30 | 0.43 ± 0.30 | 0.41 ± 0.29 | 0.35 | >0.05 | MDD=BD_I=SSD=BD_II | Ps>0.05 |
| postcentral | 0.33 ± 0.28 | 0.31 ± 0.26 | 0.33 ± 0.29 | 0.33 ± 0.28 | 0.26 | >0.05 | MDD=BD_I=SSD=BD_II | Ps>0.05 |
| precentral | 0.36 ± 0.32 | 0.30 ± 0.29 | 0.35 ± 0.31 | 0.36 ± 0.30 | 1.98 | >0.05 | BD-I=SSD=MDD=B | Ps>0.05 |

| | | | | | | | | |
|---|---|---|---|---|---|---|---|---|
| | | | | | | | D-II | |
| precuneus | 0.35 ± 0.31 | 0.32 ± 0.28 | 0.35 ± 0.29 | 0.34 ± 0.29 | 0.66 | >0.05 | BD-I=MDD=SSD=BD-II | Ps>0.05 |
| rostralanteriorcingulate | 0.46 ± 0.28 | 0.47 ± 0.29 | 0.46 ± 0.28 | 0.46 ± 0.27 | 0.15 | >0.05 | BD-II=MDD=BD-I=SSD | Ps>0.05 |
| rostralmiddlefrontal | 0.34 ± 0.33 | 0.32 ± 0.30 | 0.37 ± 0.33 | 0.35 ± 0.31 | 1.23 | >0.05 | MDD=SSD=BD-I=BD-II | Ps>0.05 |
| superiorfrontal | 0.35 ± 0.30 | 0.34 ± 0.29 | 0.40 ± 0.31 | 0.35 ± 0.28 | 2.64 | P_raw<0.05; P_adj >0.05 | MDD=BD-I=SSD=BD-II | Ps>0.05 |
| superiorparietal | 0.41 ± 0.31 | 0.36 ± 0.28 | 0.38 ± 0.29 | 0.36 ± 0.27 | 1.80 | >0.05 | BD-I=MDD=BD-II=SSD | Ps>0.05 |
| superiortemporal | 0.32 ± 0.30 | 0.29 ± 0.28 | 0.32 ± 0.29 | 0.29 ± 0.27 | 1.09 | >0.05 | BD-I=MDD=SSD=BD-II | Ps>0.05 |
| supramarginal | 0.40 ± 0.31 | 0.38 ± 0.28 | 0.41 ± 0.29 | 0.38 ± 0.29 | 0.59 | >0.05 | BD-I=MDD=SSD=BD-II | Ps>0.05 |
| temporal pole | 0.33 ± 0.24 | 0.29 ± 0.25 | 0.28 ± 0.22 | 0.31 ± 0.23 | 2.14 | >0.05 | BD-I=SSD=BD-II=MDD | Ps>0.05 |
| transversetemporal | 0.40 ± 0.29 | 0.38 ± 0.27 | 0.40 ± 0.28 | 0.36 ± 0.26 | 1.03 | >0.05 | MDD=BD-I=BD-II=SSD | Ps>0.05 |

**Table S4.** Comparisons of social psychological environment between patient groups and healthy controls

| | **Positive social environment** | **t** | $P_{FDR}$ | **Negative social environment** | **t** | $P_{FDR}$ |
|---|---|---|---|---|---|---|
| BD-I, n=254 | -0.05±0.26 | -11.84 | <0.001 | 0.18±0.39 | 15.82 | <0.001 |

| | | | | | | |
|---|---|---|---|---|---|---|
| HC, n=715 | 0.17±0.24 | | | -0.24±0.27 | | |
| BD-II, n=278 | -0.15±0.21 | -20.89 | <0.001 | 0.22±0.41 | 17.26 | <0.001 |
| HC, n=715 | 0.17±0.24 | | | -0.24±0.27 | | |
| MDD, n=304 | -0.18±0.22 | -22.63 | <0.001 | 0.17±0.43 | 15.69 | <0.001 |
| HC, n=713 | 0.17±0.24 | | | -0.24±0.27 | | |
| SSD, n=140 | -0.03±0.29 | -7.57 | <0.001 | 0.04±0.39 | 8.05 | <0.001 |
| HC, n=714 | 0.17±0.24 | | | -0.24±0.27 | | |

Note: We used propensity score matching to create demographically matched (on age and gender) control groups for each diagnosis, enabling direct comparisons of environmental factors between patient groups and their respective HCs.

**Table S5.** Comparisons of natural environment between patient groups and healthy controls

| | **$PM_{2.5}$** | **t** | **$P_{FDR}$** | **NDVI** | **t** | **$P_{FDR}$** |
|---|---|---|---|---|---|---|
| BD-I, n=251 | 35.91±5.15 | 6.51 | <0.001 | 0.45±0.04 | -4.14 | <0.001 |
| HC, n=713 | 33.61±3.69 | | | 0.46±0.03 | | |
| BD-II, n=273 | 36.20±6.98 | 5.84 | <0.001 | 0.45±0.05 | -3.37 | <0.05 |

| | | | | | | |
|---|---|---|---|---|---|---|
| HC, n=714 | 33.61±3.69 | | | 0.46±0.03 | | |
| MDD, n=292 | 35.66±6.02 | 5.39 | <0.001 | 0.45±0.05 | -3.89 | <0.001 |
| HC, n=712 | 33.61±3.69 | | | 0.46±0.03 | | |
| SSD, n=137 | 36.19±5.13 | 5.61 | <0.001 | 0.45±0.04 | -3.29 | <0.05 |
| HC, n=713 | 33.61±3.69 | | | 0.46±0.03 | | |

Note: We used propensity score matching to create demographically matched (on age and gender) control groups for each diagnosis, enabling direct comparisons of environmental factors between patient groups and their respective HCs.

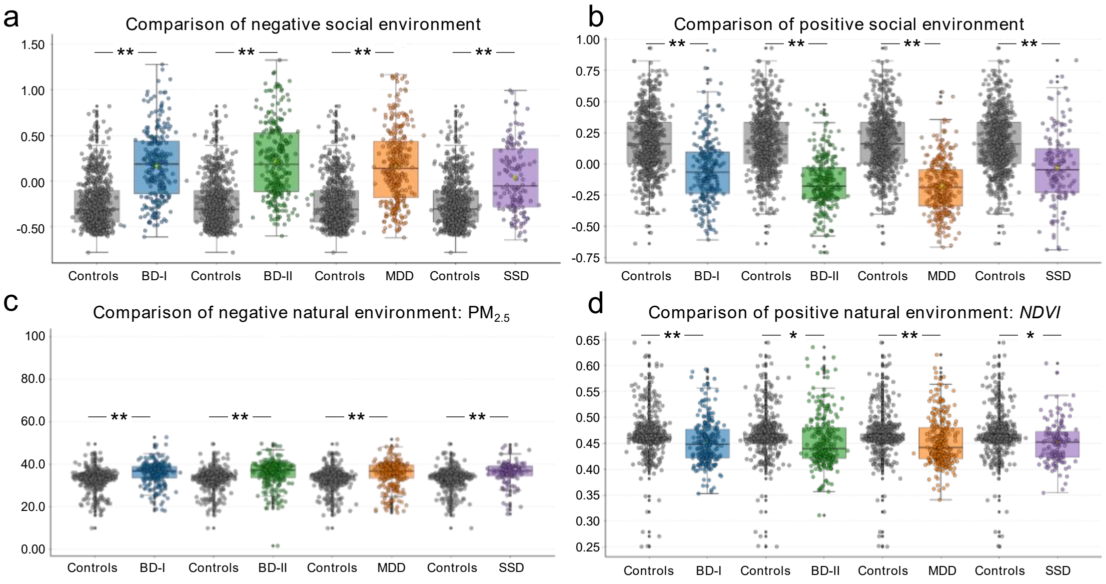


**Fig. S1 Group differences in social and natural environment factors**. (a) Negative social, (b) positive social, (c) negative natural, and (d) positive natural. All patient groups were demographically matched (age and gender) to controls using propensity score matching.

Note: Asterisks denote statistical significance: *$P_{FDR} < 0.05$, **$P_{FDR} < 0.001$. BD-I, bipolar disorder type I; BD-II, bipolar disorder type II; MDD, major depressive disorder; SSD, schizophrenia spectrum disorders.

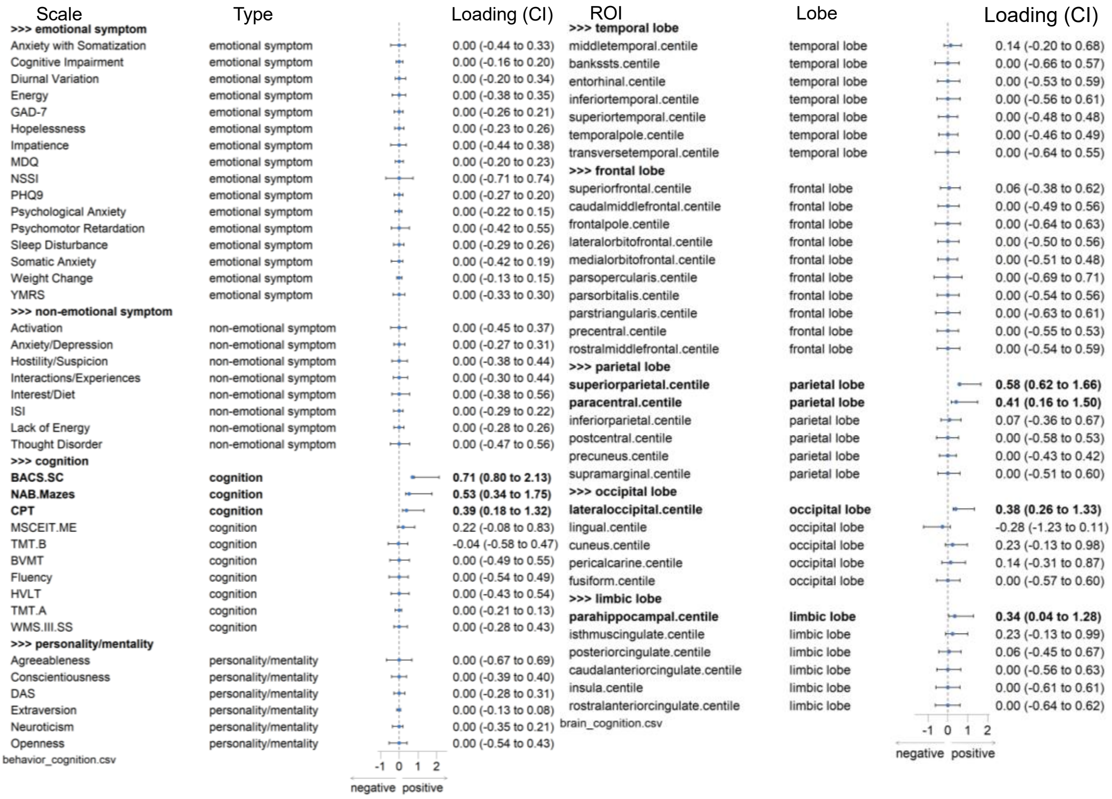


**Fig. S2 Resampling distributions for behavioral and brain features for Mode 1 in the full sample.** Features in bold are those with 95% confidence intervals not crossing zero, indicating stable contributions to the mode.

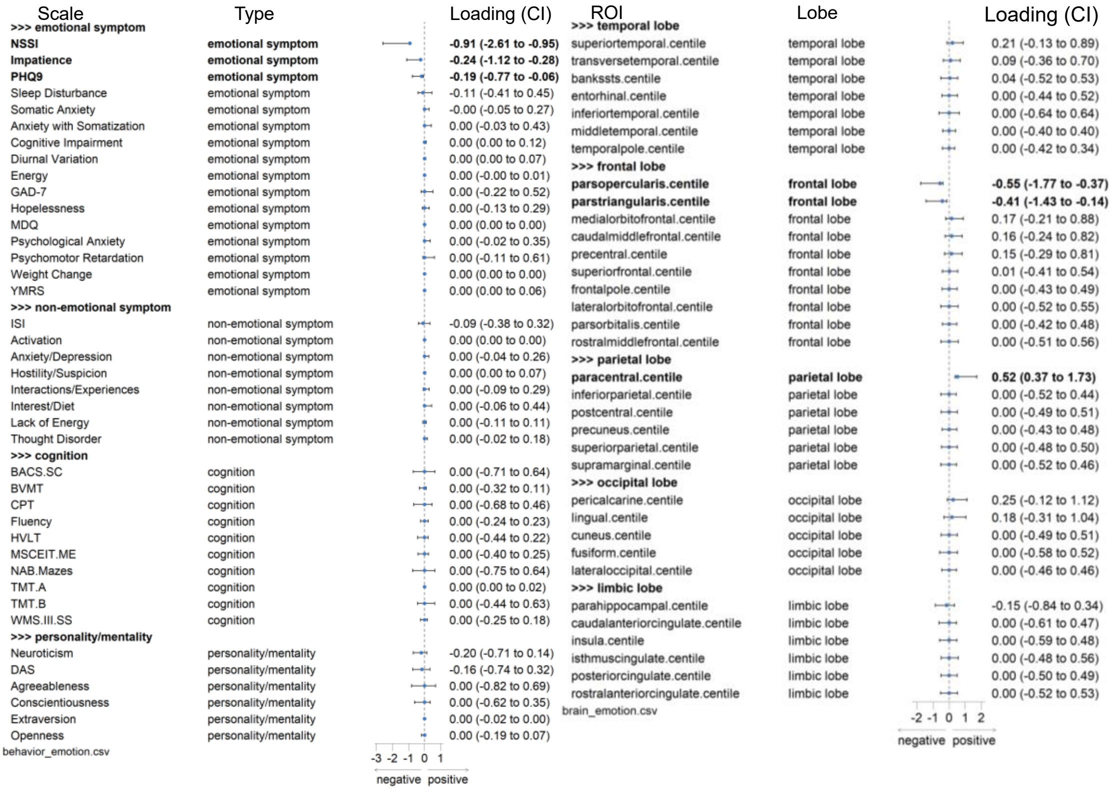


**Fig. S3 Resampling distributions for behavioral and brain features for Mode 2 in the full**

**sample.** Features in bold are those with 95% confidence intervals not crossing zero, indicating stable contributions to the mode.

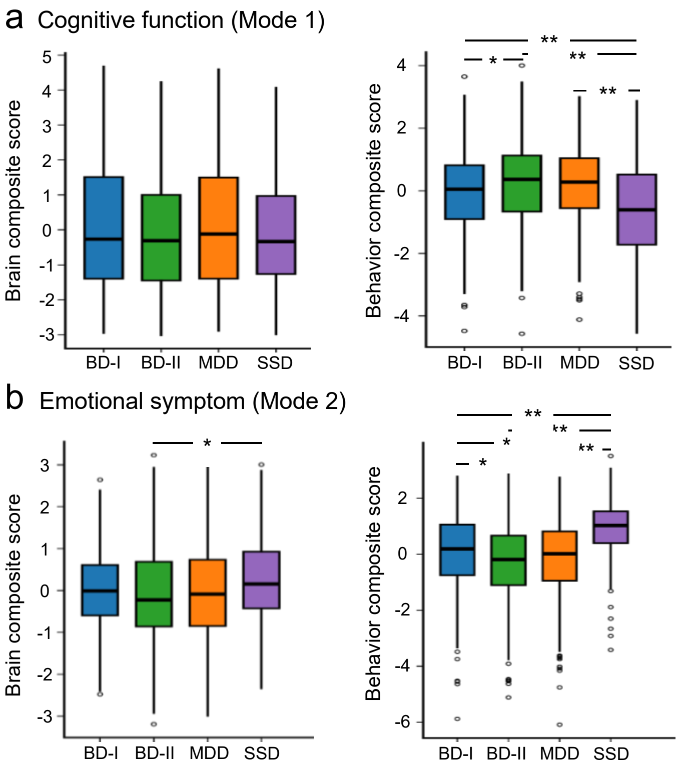


**Fig. S4 Transdiagnostic group differences in canonical brain-behavior modes.** Group comparison of brain and behavioral composite scores for Mode 1 **(a)** and Mode 2 **(b)** across the four diagnostic groups. Asterisks indicate significant between-group differences (two-sample t-tests; $*P_{FDR} < 0.05$, $**P_{FDR} < 0.001$). Abbreviations: BD-I, bipolar disorder type I; BD-II, bipolar disorder type II; MDD, major depressive disorder; SSD, schizophrenia spectrum disorders.

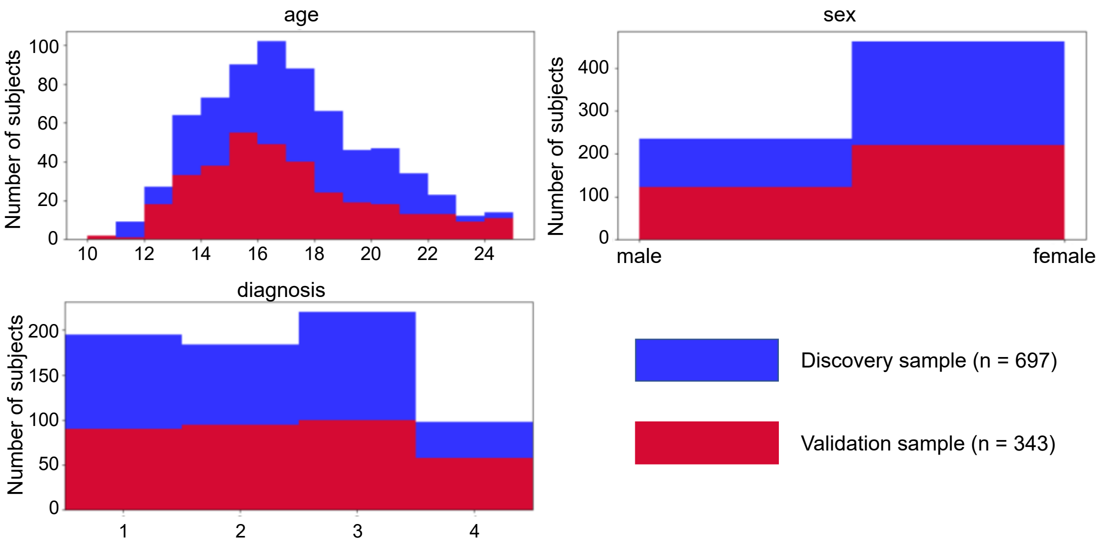


**Fig. S5 Demographic characteristics of the discovery and validation samples**. Both samples demonstrated comparable distributions in age, sex, and diagnosis.

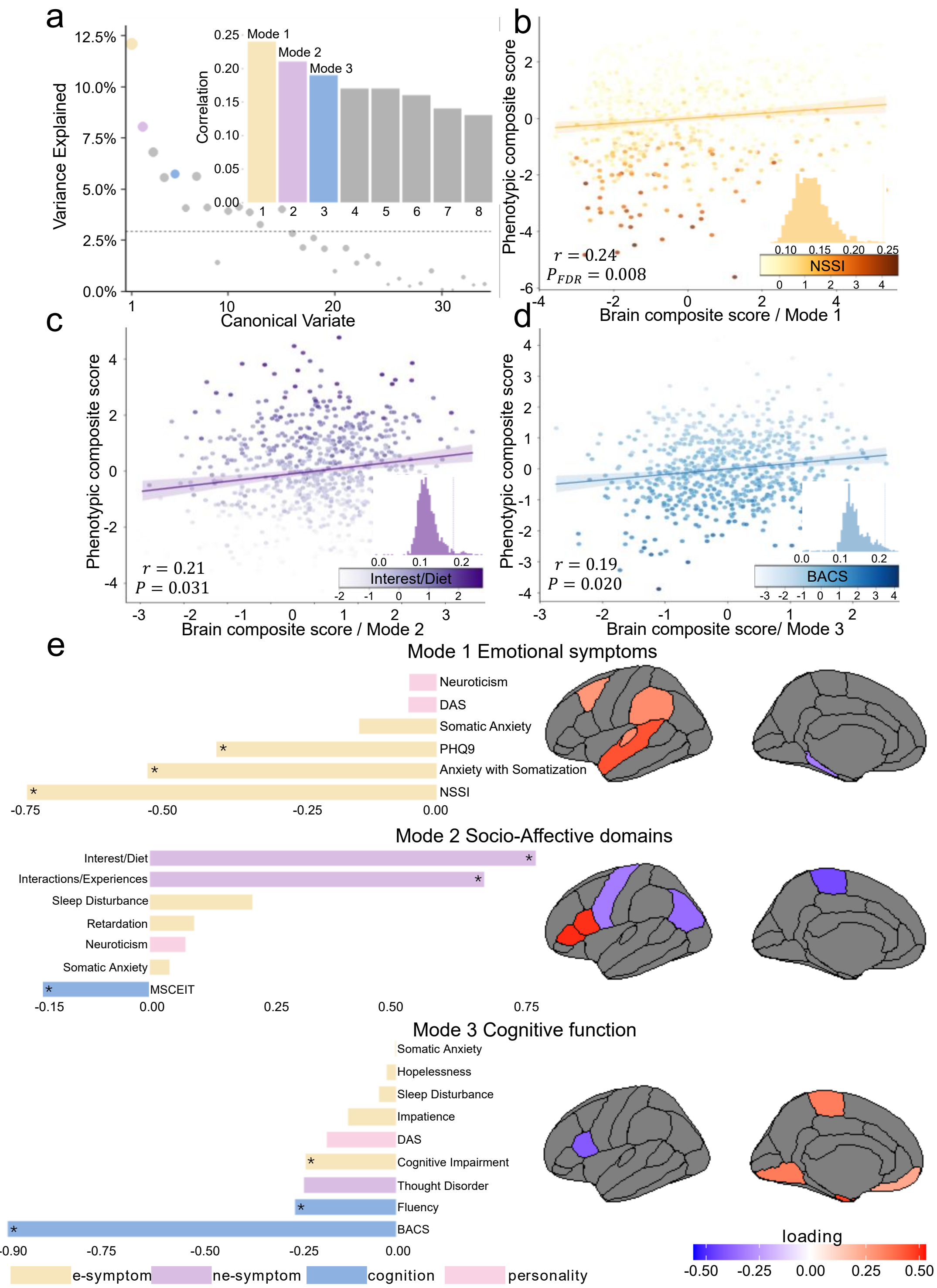


**Fig. S6 Multivariate brain-behavior relationships identified by sCCA in the discovery sample.** **a** The first 8 canonical variants ranked by explained covariance. The dashed horizontal line indicates the mean explained covariance. Statistical significance (assessed via permutation testing with FDR correction): Mode 1($P_{\mathrm{FDR}} < 0.01$), Mode 2 and Mode 3 (effects at uncorrected $P < 0.05$ threshold). **b-d** Scatter plots show the multivariate relationship between brain composite scores (x-axis) and behavior composite scores (y-axis) for Mode 1 **(b)**, Mode 2 **(c)**, and Mode 3 **(d)**. Each point represents a participant; color intensity indicates the severity of the dominant clinical behavioral feature in each mode. Insets: Null distribution of the canonical correlation derived from permutation testin. **e** Behavioral and brain loadings for Mode 1-3.

Insets: Asterisks indicate features that have significant loadings based on bootstrap resampling (95% CI $\neq 0$).

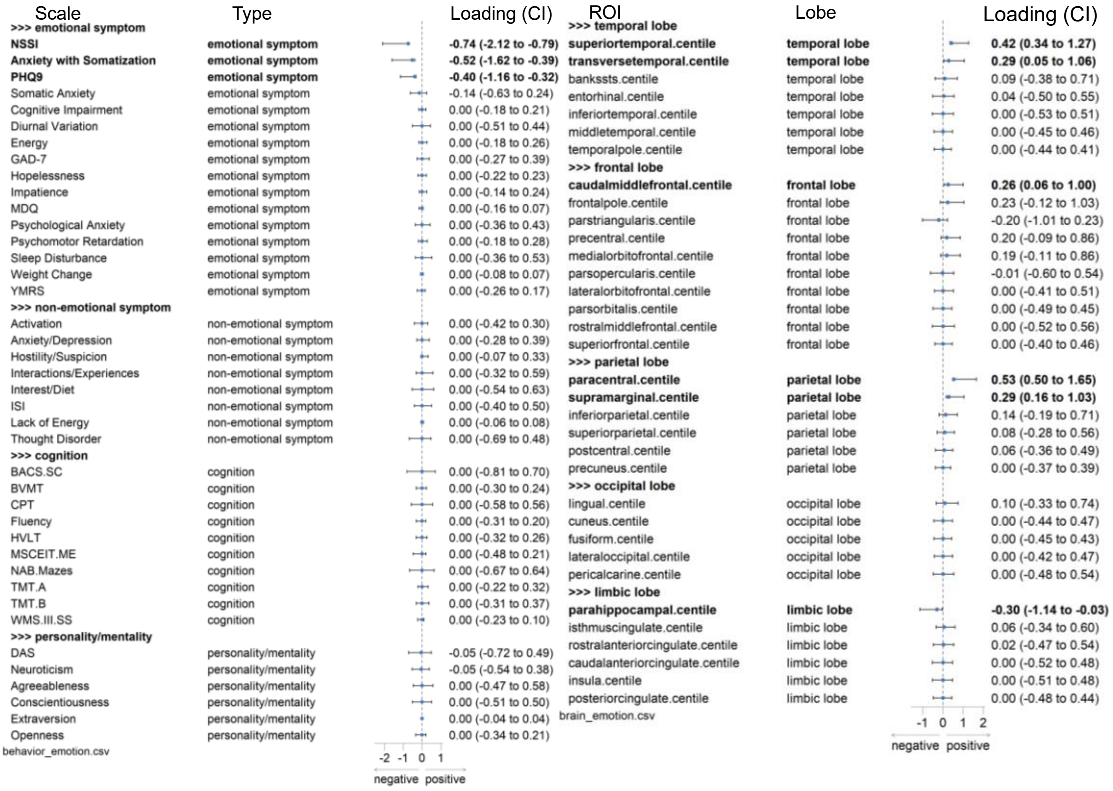


**Fig. S7 Resampling distributions of behavioral and brain features for Mode 1 in the discovery dataset.** Features in bold have 95% confidence intervals that do not cross zero, indicating their stable contributions to the mode.

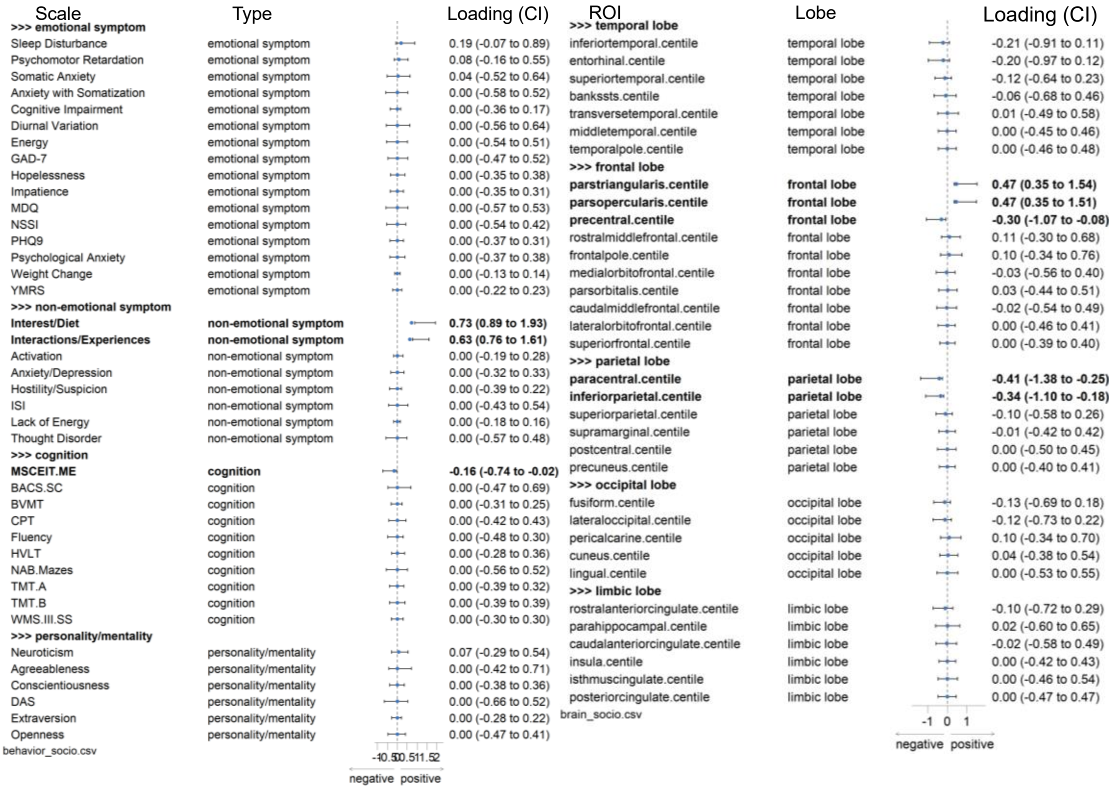


**Fig. S8 Resampling distributions of behavioral and brain features in Mode 2 in the discovery dataset.** Features in bold have 95% confidence intervals that do not cross zero, indicating their stable contributions to the mode.

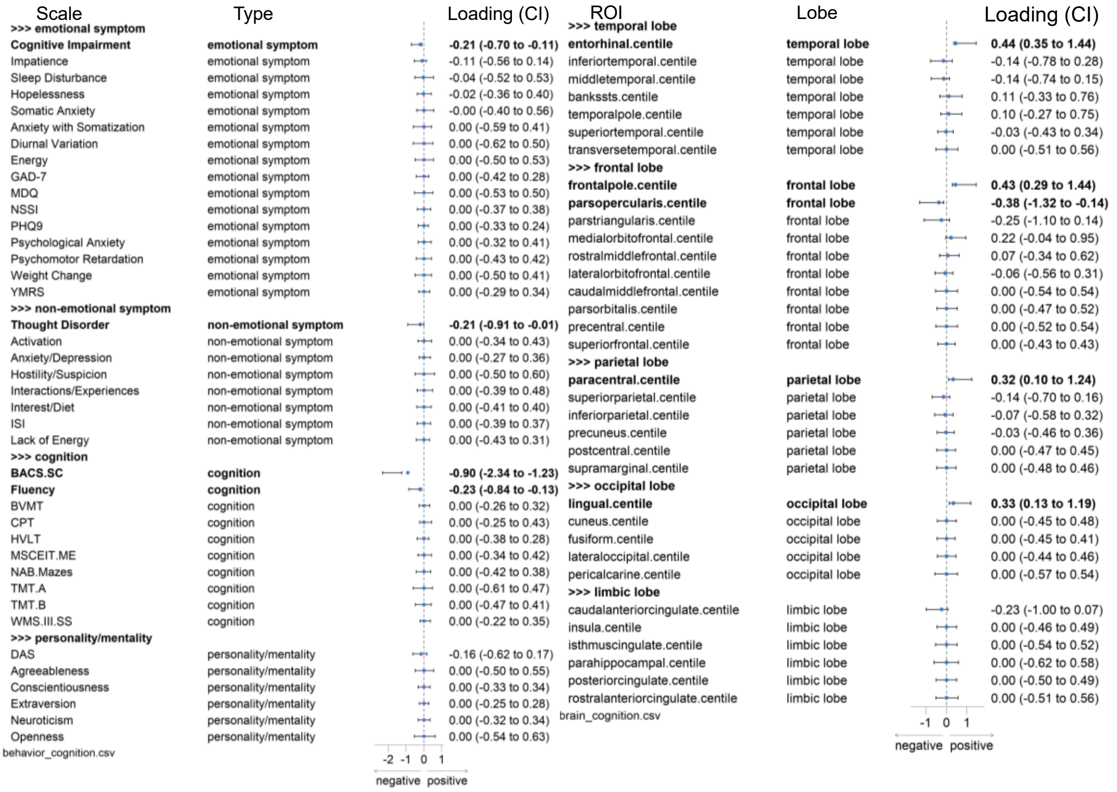


**Fig. S9 Resampling distributions of behavioral and brain features in Mode 3 in the discovery**

**dataset.** Features in bold have 95% confidence intervals that do not cross zero, indicating their stable contributions to the mode.

### Independent validation in the validation sample

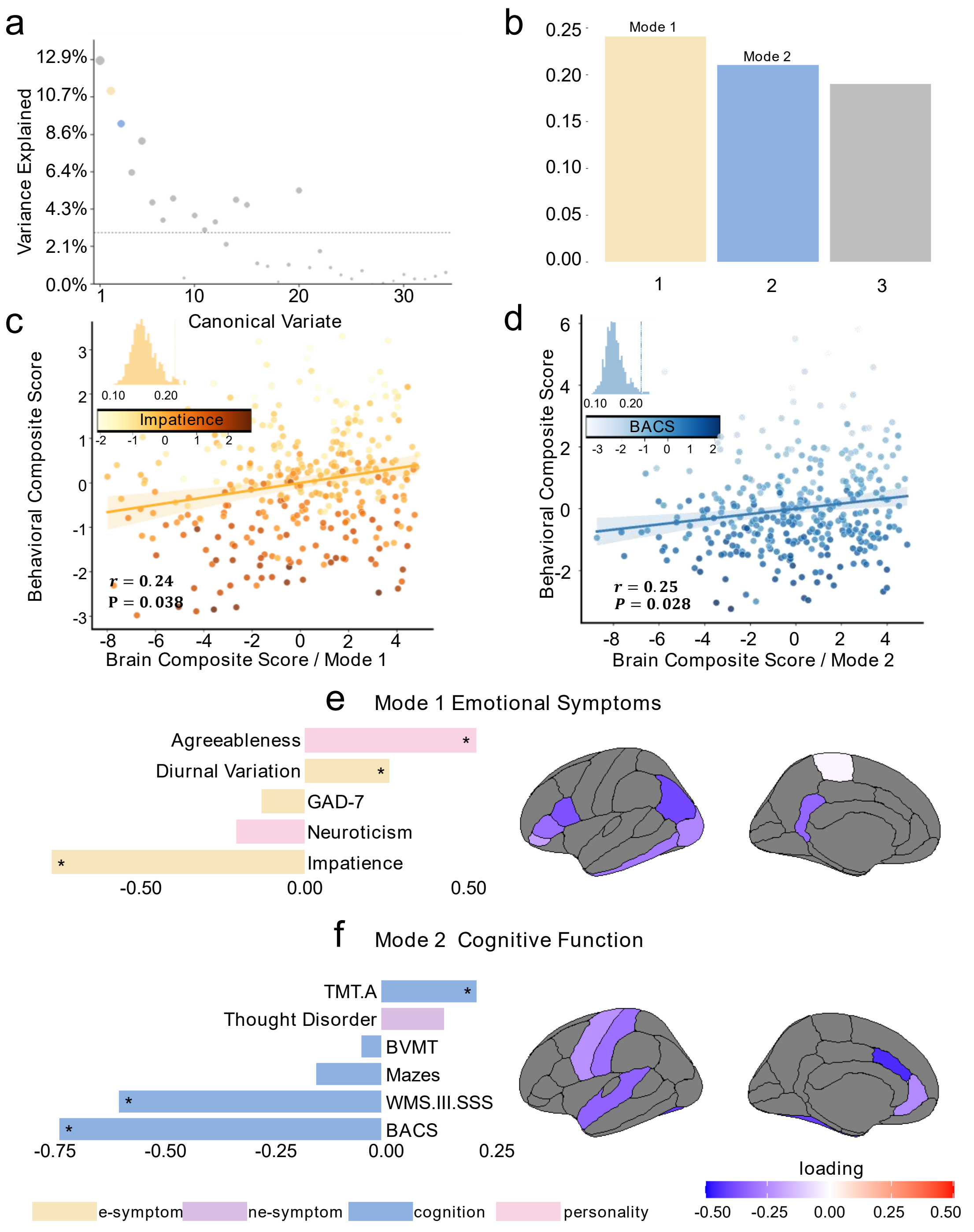


**Fig. S10 Multivariate brain-behavior relationships identified by sCCA in the validation sample. a** Explained covariance of the first three canonical modes. The dashed horizontal line indicates the mean explained covariance. **b** Statistical significance of each mode assessed by permutation testing with FDR correction. Mode 1: $P < 0.05$ (uncorrected); Mode 2: $P < 0.05$ (uncorrected). Significant modes are highlighted in bright shades. **c-d** Scatter plots of brain composite scores (x-axis) against behavior composite scores (y-axis) for Mode 1 **(c)** and Mode 2 **(d)**. Each point represents one participant; color intensity corresponds to the severity of the dominant clinical behavioral feature in each mode. **e-f:** Behavioral and brain loadings for Mode 1 (“Emotional Symptoms”) and Mode 2 (“Cognitive Function”).

Insets: Asterisks indicate features that have significant loadings based on bootstrap resampling (95% CI$\neq$0).

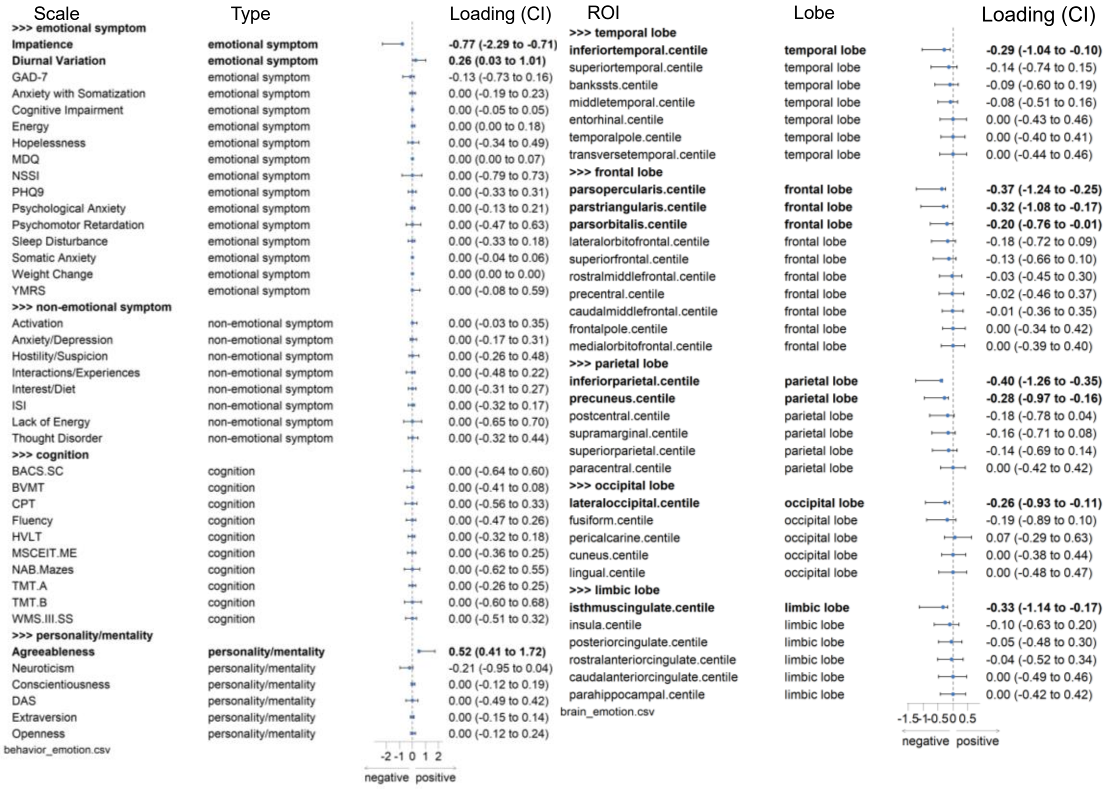


**Fig. S11 Resampling distributions of behavioral and brain features in Mode 1 in the validation dataset.** Features in bold have 95% confidence intervals that do not cross zero, indicating their stable contributions to the mode.

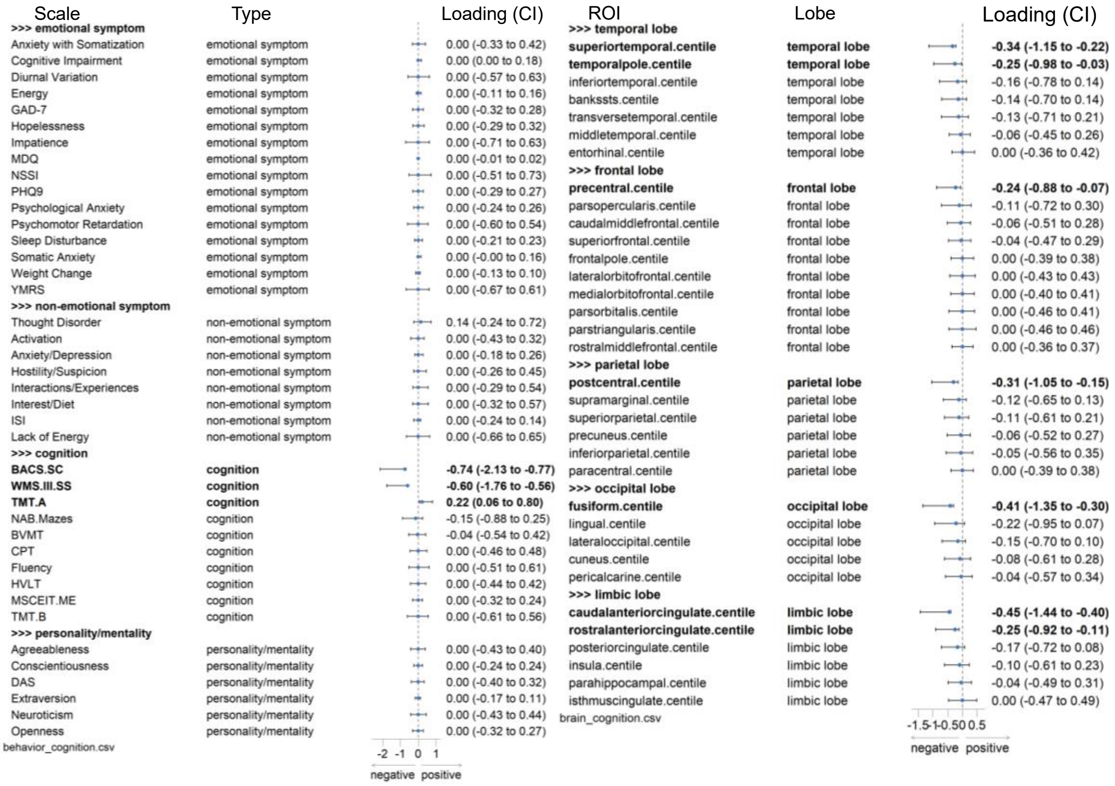


**Fig. S12 Resampling distribution of behavioral and brain features in Mode 2 in the validation dataset.** Features in bold have 95% confidence intervals that do not cross zero, indicating their stable contributions to the mode.

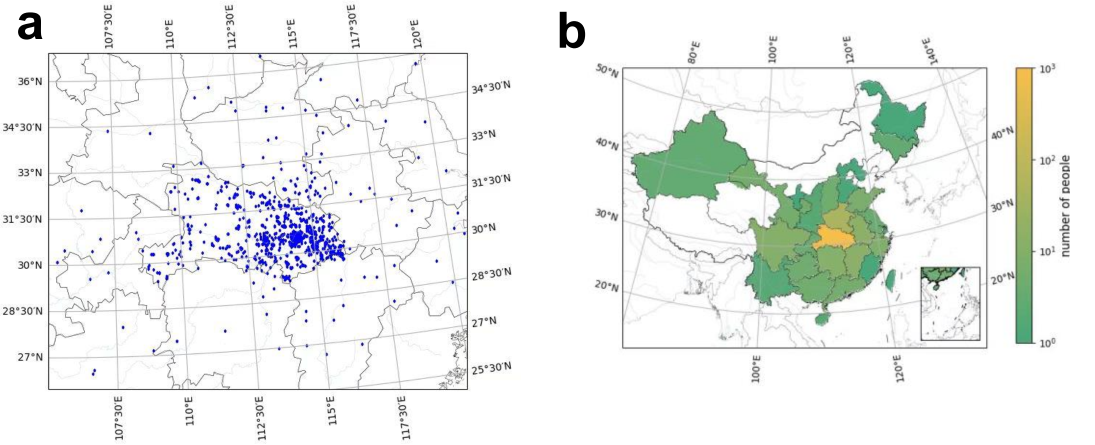


**Fig. S13 Geographical distribution of sampling locations.** The base map was obtained from the Standard Map Service of the Ministry of Natural Resources of China (Drawing Review No. GS(2016)1600) and is unmodified in its cartographic representation.

### Supplementary Methods

#### Assessment of emotion-related and non-emotion-related clinical symptoms

Initially developed by Zheng Ying et al., and later revised by Feng Yu [1], this scale measures the frequency and severity of 18 types of NSSI behaviors, including cutting, burns/scalds, stabbing, and ligature injuries. It employs a 4-point scoring system: frequency scores of 0, 1, 2, and 3 indicate "0 times", "1 times", "2-4 times" and "5 times", respectively, while severity scores of 0, 1, 2, and 3 correspond to "none", "mild", "moderate", and "severe". "Mild" denotes minor harm that resolves without intervention; "moderate" indicates injury requiring treatment with slight limitation in individual functional activity; and "severe" refers to physical damage necessitating hospitalization. The overall self-injury severity (total score) is comprehensively assessed by multiplying the frequency of NSSI behaviors by the degree of physical harm. This scale demonstrates good reliability and validity and has been widely used in the assessment of adolescent self-harm in China.

**Table S6.** Description of Adolescent Non-suicidal Self-injury Questionnaire

| Behavior in Your Past Life | Number of Occurrences (times) | | | |
|---|---|---|---|---|
| | 0 | 1 | 2-4 | 5 |
| | The degree of harm to the body | | | |
| | none | mild | moderate | severe |
| 1-Intentionally cut own skin with glass, knife, etc. | | | | |
| 2-Intentionally picked at wounds to prevent healing. | | | | |
| 3-Intentionally burned/scalded own skin with cigarettes, lighters, or other objects. | | | | |
| 4-Intentionally carved words or patterns on the body (excluding tattoos for aesthetic purposes). | | | | |
| 5-Intentionally scraped own skin until bleeding. | | | | |
| 6-Intentionally pierced skin or inserted objects under nails | | | | |
| 7-Intentionally banged head against objects, resulting in bruising. | | | | |
| 8-Intentionally pulled out own hair. | | | | |
| 9-Intentionally punched walls, glass, or other hard objects. | | | | |
| 10-Intentionally scratched oneself violently, causing scars or bleeding. | | | | |
| 11-Intentionally pierced body parts with needles, nails, or other objects to draw blood. | | | | |
| 12-Intentionally rubbed skin until bleeding. | | | | |
| 13-Intentionally hit oneself, resulting in bruising. | | | | |
| 14-Intentionally tied ropes or other objects around wrists or other body parts to constrict. | | | | |

| |
|---|
| 15-Intentionally allowed others to hit or bite oneself to cause bodily harm. |
| 16-Intentionally exposed oneself to electric shock in non-life-threatening situations. |
| 17-Intentionally bit oneself until the skin broke. |
| 18-Intentionally set fire to hands or touched flames. |

**Table S7.** Description of emotion-related clinical symptoms

| Scales | Item Description | Factors |
|---|---|---|
| HCL-32, two factors | 1-I need less sleep. | energy |
| | 2-I feel more energetic and more active. | energy |
| | 3-I am more self-confident. | energy |
| | 4-I enjoy my work more. | energy |
| | 5-I am more sociable (make more phone calls, go out more). | energy |
| | 6-I want to travel and/or do travel more. | energy |
| | 7-I tend to drive faster or take more risks when driving. | impatience |
| | 8-I spend more money/too much money. | impatience |
| | 9-I take more risks in my daily life (in my work and/or other activities). | impatience |
| | 10-I am physically more active (sport, etc.). | energy |
| | 11-I plan more activities or projects. | energy |
| | 12-I have more ideas, I am more creative. | energy |
| | 13-I am less shy or inhibited. | energy |
| | 14-I wear more colourful and more extravagant clothes/make-up. | energy |
| | 15-I want to meet or actually do meet more people. | energy |
| | 16-I am more interested in sex, and/or have increased sexual desire. | energy |
| | 17-I want to attract the attention of more members of the opposite sex. | energy |
| | 18-I talk more. | energy |
| | 19-I think faster. | energy |
| | 20-I make more jokes or puns when I am talking. | energy |
| | 21-I am more easily distracted. | impatience |
| | 22-I engage in lots of new things. | energy |
| | 23-My thoughts jump from one topic to another. | impatience |
| | 24-I do things more quickly and/or more easily. | energy |
| | 25-I am more impatient and/or get irritable more easily. | impatience |
| | 26-I can be exhausting for others. | impatience |
| | 27-I get into more quarrels. | impatience |

| | | |
|---|---|---|
| | 28-My mood is higher, more optimistic. | energy |
| | 29-I drink more coffee or tea. | impatience |
| | 30-I smoke more cigarettes. | impatience |
| | 31-I drink more alcohol. | impatience |
| | 32-I take more drugs (sedatives, anxiolytics, stimulants, etc.). | impatience |
| HAMA, two factors | 1-anxious mood | psychic anxiety |
| | 2-tension | psychic anxiety |
| | 3-fears | psychic anxiety |
| | 4-insomnia | psychic anxiety |
| | 5-cognition | psychic anxiety |
| | 6-depressed mood | psychic anxiety |
| | 7-somatic (muscular) | somatic anxiety |
| | 8-somatic (sensory) | somatic anxiety |
| | 9-cardiovascular symptoms | somatic anxiety |
| | 10-respiratory symptoms | somatic anxiety |
| | 11-gastrointestinal symptoms | somatic anxiety |
| | 12-genitourinary symptoms | somatic anxiety |
| | 13-autonomic symptoms | somatic anxiety |
| | 14-behavior at interview | psychic anxiety |
| HAMD-24, seven factors | 1-depressed mood | retardation |
| | 2-feelings of guilt | cognitive disturbance |
| | 3-suicide | cognitive disturbance |
| | 4-insomnia early | sleep disturbance |
| | 5-insomnia middle | sleep disturbance |
| | 6-insomnia late | sleep disturbance |
| | 7-work and activities | retardation |
| | 8-retardation: psychomotor | retardation |
| | 9-agitation | cognitive disturbance |
| | 10-anxiety psychic | anxiety/somatization |
| | 11-anxiety somatic | anxiety/somatization |
| | 12-somatic symptoms gastrointestinal | anxiety/somatization |
| | 13-somatic symptoms general | anxiety/somatization |
| | 14-genital symptoms | retardation |
| | 15-hypochondriasis | anxiety/somatization |
| | 16-loss of weight | weight |
| | 17-insight | anxiety/somatization |
| | 18-diurnal variation | diurnal variation |
| | 19-depersonalization and derealization | cognitive disturbance |
| | 20-paranoid symptoms | cognitive disturbance |
| | 21-obsessional and compulsive symptoms | cognitive disturbance |
| | 22-helplessness | hopelessness |
| | 23-hopelessness | hopelessness |

| | | |
|---|---|---|
| | 24-worthlessness | hopelessness |
| GAD-7 | 7 items | total score |
| PHQ-9 | 9 items | total score |
| MDQ | 14 items | total score |
| NSSI | 36 items | total score |
| YMRS | 11 items | total score |

Note: HCL-32, Hypomania Checklist; HAMA, Hamilton Anxiety Scale; HAMD-24, Hamilton Depression Rating Scale-24; GAD-7, Generalized Anxiety Disorder-7; PHQ-9, Patient Health Questionnaire-9; MDQ, Mood Disorder Questionnaire; NSSI, Adolescent Non-suicidal Self-injury Questionnaire; YMRS, Young Mania Rating Scale.

**Table S8.** Description of non-emotion-related clinical symptoms

| | Item Description | Factors |
|---|---|---|
| BPRS, five factors | 1-somatic concern | anxiety-depression |
| | 2-anxiety | anxiety-depression |
| | 3-emotional withdrawal | anergia |
| | 4-conceptual disorganization | thought disturbance |
| | 5-guilt feelings | anxiety-depression |
| | 6-tension | activation |
| | 7-mannerisms and posturing | activation |
| | 8-grandiosity | thought disturbance |
| | 9-depressive mood | anxiety-depression |
| | 10-hostility | hostility-suspicion |
| | 11-suspiciousness | hostility-suspicion |
| | 12-hallucinatory behavior | thought disturbance |
| | 13-motor retardation | anergia |
| | 14-uncooperativeness | hostility-suspicion |
| | 15-unusual thought content | thought disturbance |
| | 16-blunted affect | anergia |
| | 17-excitement | activation |
| | 18-disorientation | anergia |
| SHAPS, two factors | 1- I would enjoy my favorite television or radio program. | social/experiential pleasure |
| | 2- I would enjoy being with my family or close friends. | social/experiential pleasure |
| | 3- I would find pleasure in my hobbies and pastimes. | interest/diet |
| | 4- I would be able to enjoy my favorite meal. | interest/diet |
| | 5- I would enjoy a warm bath or refreshing shower. | interest/diet |
| | 6- I would find pleasure in the scent of flowers or the | interest/diet |

| | | |
|---|---|---|
| | smell of a fresh air or freshly baked bread. | |
| | 7- I would feel pleased when I see a friendly face. | interest/diet |
| | 8- I would enjoy looking smart or well-groomed when I have made an effort to do so. | interest/diet |
| | 9- I would enjoy reading a book, magazine or newspaper. | interest/diet |
| | 10- I would find pleasure in a cup of tea, coffee or my favorite drink. | interest/diet |
| | 11- I would feel pleased in the little things, like a sunny day or a phone call from a friend. | social/experiential pleasure |
| | 12- I would appreciate a beautiful landscape or view. | social/experiential pleasure |
| | 13- I would feel pleased when I help others. | social/experiential pleasure |
| | 14- I would be pleased to receive compliments or praise. | social/experiential pleasure |
| ISI | 5 items | total score |

Note: BPRS, Brief Psychiatric Rating Scale; SHAPS, Snaith-Hamilton Pleasure Scale; ISI, Insomnia Severity Index.

### Assessment of cognitive function: MATRICS Consensus Cognitive Battery

**Table S9.** Description of MATRICS Consensus Cognitive Battery

| Subtests | Cognitive domains |
|---|---|
| Trail Making Test-Part A (TMT-A) | speed of processing |
| Trail Making Test-Part B (TMT-B) | speed of processing |
| Symbol coding subtest from the Brief Assessment of Cognition in Schizophrenia (BACS-SC) | speed of processing |
| Hopkins Verbal Learning Test-Revised (HVLT-R) | verbal learning |
| Spatial Span subtest from the Wechsler Memory Scale-III (WMS-III. SS) | working memory |
| Mazes subtest from the Neuropsychological Assessment Battery (NAB Mazes) | reasoning and problem solving |
| Brief Visuospatial Memory Test-Revised (BVMT) | visual learning |
| Verbal Fluency (animals) | speed of processing |
| Continuous Performance Test-Identical Pairs (CPT-IP) | attention/vigilance |
| Managing Emotions component of the Mayer-Salovey-Caruso Emotional Intelligence Test (MSCEIT) | social cognition |

### Assessment of personality and maladaptive attitudes/beliefs

**Table S10.** Description of personality and maladaptive attitudes/beliefs

| | Item Description | Factors |
|---|---|---|
| NEO, five factors | 1-I am not someone who worries easily. | neuroticism |
| | 2-I like to have a lot of people around me. | extraversion |
| | 3-I enjoy concentrating on fantasy or daydream and exploring all its possibilities, letting it grow and develop. | openness |
| | 4-I try to be courteous to everyone I meet. | agreeableness |
| | 5-I keep my belongings neat and clean. | conscientiousness |
| | 6-Sometimes I feel angry and full of resentment. | neuroticism |
| | 7-I laugh easily. | extraversion |
| | 8-I think it's interesting to learn and develop new hobbies. | openness |
| | 9-Sometimes I use different tactics, such as threats or flattery, to persuade others to do what I want. | agreeableness |
| | 10-I'm pretty good about pacing myself so as to get things done on time. | conscientiousness |
| | 11-When I'm under a great deal of stress, sometimes I feel like I'm going to pieces. | neuroticism |
| | 12- I prefer jobs that allow me to work independently without being disturbed by others. | extraversion |
| | 13-I am intrigued by the patterns I find in art and nature. | openness |
| | 14-Some people think I'm selfish and egotistical. | agreeableness |
| | 15-I often come into situations without being fully prepared. | conscientiousness |
| | 16-I rarely feel lonely or blue. | neuroticism |
| | 17-I really enjoy chatting with others. | extraversion |
| | 18-I believe letting students hear controversial speakers can only confuse and mislead them. | openness |
| | 19-If someone starts a fight, I'm ready to fight back. | agreeableness |
| | 20-I try to perform all the tasks assigned to me conscientiously. | conscientiousness |
| | 21-I often feel tense and jittery. | neuroticism |
| | 22-I enjoy being involved in intense activities. | extraversion |
| | 23-Poetry has little or no effect on me. | openness |
| | 24-I'm better than most people, and I know it. | agreeableness |
| | 25-I have clear goals and can work towards them in a systematic manner. | conscientiousness |
| | 26-Sometimes I feel completely worthless. | neuroticism |
| | 27-I usually avoid crowded situations. | extraversion |
| | 28-I would have difficulty just letting my mind wander | openness |

| | | |
|---|---|---|
| | without control or guidance. | |
| | 29-When I've been insulted, I just try to forgive and forget. | agreeableness |
| | 30-I waste a lot of time before settling down to work. | conscientiousness |
| | 31-I rarely feel fearful or anxious. | neuroticism |
| | 32- I often feel energetic, as if I'm full of energy. | extraversion |
| | 33-I seldom notice the moods or feelings that different environments produce | openness |
| | 34-I tend to assume the best about people. | agreeableness |
| | 35-I work hard to accomplish my goals. | conscientiousness |
| | 36-I often get angry at the way people treat me. | neuroticism |
| | 37-I am an optimistic and cheerful person. | extraversion |
| | 38-I experience a wide range of emotions or feelings. | openness |
| | 39-Some people think of me as cold and calculating. | agreeableness |
| | 40-When I make a commitment, I can always be counted on to follow through. | conscientiousness |
| | 41-Too often, when things go wrong, I get discouraged and feel like giving up | neuroticism |
| | 42-I don't really enjoy chatting with people and rarely get much pleasure from it. | extraversion |
| | 43-Sometimes when I am reading poetry or looking at a work of art, I feel a chill or a wave of excitement. | openness |
| | 44- I have no sympathy for beggars. | agreeableness |
| | 45-Sometimes I'm not as dependable or reliable as I should be. | conscientiousness |
| | 46-I am seldom sad or depressed. | neuroticism |
| | 47-My pace of life is very fast. | extraversion |
| | 48-I have little interest in speculating on the nature of the universe or the human condition. | openness |
| | 49-I generally try to be thoughtful and considerate. | agreeableness |
| | 50-I am a productive person who always gets the job done. | conscientiousness |
| | 51-I often feel helpless and want someone else to solve my problems. | neuroticism |
| | 52-I am a very active and energetic person. | extraversion |
| | 53- I have a lot of intellectual curiosity. | openness |
| | 54-If I don't like people, I let them know it. | agreeableness |
| | 55-I never seem to be able to get organized. | conscientiousness |
| | 56-At times I have been so ashamed I just wanted to hide. | neuroticism |

| | | |
|---|---|---|
| | 57-I prefer to do things on my own rather than lead or direct others. | extraversion |
| | 58-I often enjoy playing with theories or abstract ideas. | openness |
| | 59-If necessary, I am willing to manipulate people to get what I want. | agreeableness |
| | 60-I strive for excellence in everything I do. | conscientiousness |
| DAS | 40 items | total score |

Note: NEO, NEO Personality Inventory; DAS, Dysfunctional Attitudes Scale.

### Assessment of positive and negative social psychological environment

**Table S11.** Description of social psychological environment: parental parenting style

| | Item Description | Factors of maternal parenting style | Factors of paternal parenting style |
|---|---|---|---|
| Memories of Parental Rearing Behaviour: four factors in maternal parenting style; five factors in parental parenting style. | 1-I feel that my parents interfere with everything I do. | Overcontrol/Overprotection | Overcontrol |
| | 2-I can tell from my parents' words and expressions that they like me. | Emotional Warmth | Emotional Warmth |
| | 3-My parents are always overly worried about my health. | Overcontrol/Overprotection | Overprotection |
| | 4-Even for minor mistakes, my parents punish me. | - | Punishment |
| | 5-My parents always try to subtly influence me to become outstanding. | Emotional Warmth | Emotional Warmth |
| | 6-I feel that my parents allow me to have unique traits in certain aspects. | Emotional Warmth | Emotional Warmth |
| | 7-The punishment from my parents is fair and appropriate. | Emotional Warmth | Emotional Warmth |
| | 8-I feel that my parents are very strict with me. | - | Overcontrol |
| | 9-My parents always control what I should wear or how I should look. | Overcontrol/Overprotection | Overcontrol |
| | 10-My parents don't allow me to do things that other children are allowed to do because they're afraid something might happen to me. | Overcontrol/Overprotection | Overprotection |
| | 11-When I was young, my parents | Punishment | Punishment |

| | | | |
|---|---|---|---|
| | hit or scolded me in front of others. | | |
| | 12-My parents always monitor what I do at night. | Overcontrol/Overprotection | Overcontrol |
| | 13-When I'm upset, I can feel my parents trying to encourage and comfort me. | Emotional Warmth | Emotional Warmth |
| | 14-I can feel that my parents like me. | Emotional Warmth | Emotional Warmth |
| | 15-My parents' punishment is always more severe than I deserve. | Punishment | Punishment |
| | 16-If I don't follow instructions at home, my parents get angry. | - | Punishment |
| | 17-If I do something wrong, my parents make me feel guilty or remorseful by acting hurt. | Overcontrol/Overprotection | - |
| | 18-I find it difficult to get close to my parents. | - | Emotional Warmth |
| | 19-My parents have embarrassed me by complaining about things I said or did in front of others. | - | Permissiveness |
| | 20-My parents are stingy when it comes to meeting my needs. | Permissiveness | Permissiveness |
| | 21-My parents often care a lot about the grades I get. | Overcontrol/Overprotection | - |
| | 22-When facing a difficult task, I can feel support from my parents. | Emotional Warmth | Emotional Warmth |
| | 23-At home, I am often made the "scapegoat" or "black sheep." | Permissiveness | - |
| | 24-My parents always criticize the friends I like. | Overcontrol/Overprotection | Overcontrol |
| | 25-My parents always think that their unhappiness is caused by me. | Permissiveness | Permissiveness |
| | 26-My parents always try to encourage me to be the best. | Emotional Warmth | Emotional Warmth |
| | 27-My parents show me that they love me. | Emotional Warmth | Emotional Warmth |
| | 28-My parents trust me and allow me to complete certain things on my own. | Emotional Warmth | Emotional Warmth |
| | 29-I feel that my parents respect my opinions. | Emotional Warmth | Emotional Warmth |
| | 30-I feel that my parents enjoy spending time with me. | Emotional Warmth | Emotional Warmth |
| | 31-I feel that my parents are stingy | Permissiveness | Permissiveness |

| | | | |
|---|---|---|---|
| | and mean with me. | | |
| | 32-My parents often say things like, "It would break my heart if you did this." | Overcontrol/Overprotection | Permissiveness |
| | 33-My parents require me to explain what I'm doing when I come home. | Overcontrol/Overprotection | Overcontrol |
| | 34-I feel that my parents try hard to make my youth meaningful and colourful (e.g., buying me books, sending me to summer camp or clubs). | Emotional Warmth | Emotional Warmth |
| | 35-My parents often say things like, "Is this the thanks we get for all we've done for you?" | Permissiveness | - |
| | 36-My parents often use the excuse "we can't spoil you" to deny my requests. | Permissiveness | Overprotection |
| | 37-If I don't meet my parents' expectations, I feel very guilty. | - | Overprotection |
| | 38-I feel that my parents have high expectations for my academic performance, sports, or similar activities. | Overcontrol/Overprotection | - |
| | 39-I can seek comfort from my parents when I feel sad. | Emotional Warmth | Emotional Warmth |
| | 40-My parents have punished me for no reason. | Punishment | Punishment |
| | 41-My parents allow me to do things that my friends are allowed to do. | Emotional Warmth | - |
| | 42-My parents often tell me they don't like my behaviour at home. | Permissiveness | Permissiveness |
| | 43-During meals, my parents urge or force me to eat more. | - | Overprotection |
| | 44-My parents often criticize me in front of others as lazy and useless. | Permissiveness | - |
| | 45-My parents often pay attention to what kind of friends I associate with. | Overcontrol/Overprotection | Overcontrol |
| | 46-My parents let me develop naturally. | Overcontrol/Overprotection | Overcontrol |
| | 47-My parents are often rude to me. | Punishment | Punishment |

| | | | |
|---|---|---|---|
| | 48-Sometimes my parents punish me harshly over trivial matters. | Punishment | Punishment |
| | 49-My parents have hit me for no reason. | Punishment | Punishment |
| | 50-My parents usually take part in my hobby activities. | Emotional Warmth | Emotional Warmth |
| | 51-I often get beaten by my parents. | Punishment | Punishment |
| | 52-My parents often allow me to go places I like without being overly worried. | Overcontrol/Overprotection | Overcontrol |
| | 53-My parents have strict rules about what I can and cannot do, and they never compromise. | Overcontrol/Overprotection | Overcontrol |
| | 54-My parents often treat me in ways that embarrass me. | Punishment | Punishment |
| | 55-I think my parents' worry that something might happen to me is exaggerated. | Overcontrol/Overprotection | Overprotection |
| | 56-There is a warm, caring, and affectionate relationship between me and my parents. | Emotional Warmth | Emotional Warmth |
| | 57-My parents tolerate opinions that differ from theirs. | Emotional Warmth | Emotional Warmth |
| | 58-My parents often get angry at me for no apparent reason. | Punishment | Punishment |
| | 59-When I succeed at something, I feel that my parents are proud of me. | Emotional Warmth | - |
| | 60-My parents often hug me. | - | Emotional Warmth |

Note: "-" indicates "not scored".

**Table S12.** Description of Social Support Rating Scale

| | Item Description | Options | Factors |
|---|---|---|---|
| Social Support Rating Scale: three | 1-How many close friends do you have who can offer you support and help? | (1)None; (2)1-2; (3)3-5; (4) 6 or more | Subjective Support |
| | 2-In the past year, you have: (choose only one) | (1)Lived far from family and lived alone; (2) Frequently changed residence and mostly lived with strangers; (3)Lived with classmates, colleagues, or friends; (4)Lived with | Objective Support |

| | | | |
|---|---|---|---|
| factors | | family | |
| | 3-You and your neighbors: (choose only one) | (1)Never care about each other, only nod hello; (2)Might show some concern if there is difficulty;(3)Some neighbors care about you;(4) Most neighbors care about you | Subjective Support |
| | 4-You and your colleagues/classmates/friends: (choose only one) | (1)Never care about each other, only nod hello;(2)Might show some concern if there is difficulty;(3)Some colleagues/classmates/friends care about you;(4)Most colleagues/classmates/friends care about you. | Subjective Support |
| | 5-Support and care received from family members (check " √ " in the appropriate box) | Family Member; None; Very Little; Moderate; Full Support | Subjective Support |
| | | A. Spouse/partner;<br>B. Parents;<br>C. Children;<br>D. Siblings;<br>E. Other (e.g., in-laws) | |
| | 6- In the past, when you encountered an emergency, from whom did you receive financial support or practical help? | (1) No source<br>(2) The following sources (multiple choices allowed):<br>A. Spouse/partner<br>B. Other family members<br>C. Relatives<br>D. Friends<br>E. Colleagues<br>F. Work unit<br>G. Official or semi-official organizations (e.g., political party, union)<br>H.Non-governmental organizations (e.g., religious, social groups)<br>I. Other (please specify): | Objective Support |
| | 7- In the past, when you encountered an emergency, from whom did you receive comfort and concern? | (1) No source<br>(2) The following sources (multiple choices allowed):<br>A. Spouse/partner<br>B. Other family members<br>C. Friends<br>D. Relatives<br>E. Colleagues<br>F. Work unit | Objective Support |

| | | | |
|---|---|---|---|
| | | G. Official or semi-official organizations (e.g., political party, union)<br>H.Non-governmental organizations (e.g., religious, social groups)<br>I. Other (please specify): | |
| | 8-When you are upset, how do you confide in others? (choose only one) | (1) Never confide in anyone<br>(2) Only confide in 1-2 very close people<br>(3) Will tell if friends ask actively<br>(4) Actively share your troubles to gain support and understanding | Utilization Degree of Social Support |
| | 9-When facing difficulties, how do you seek help? (choose only one) | (1) Rely only on yourself, do not accept help from others<br>(2) Rarely ask others for help<br>(3) Sometimes ask others for help<br>(4) Often seek help from family, relatives, friends, or organizations when in difficulty | Utilization Degree of Social Support |
| | 10- Regarding group activities (e.g., political parties, religious organizations, unions, student associations): (choose only one) | (1) Never participate<br>(2) Occasionally participate<br>(3) Often participate<br>(4)Participate actively and engage enthusiastically | Utilization Degree of Social Support |

**Table S13.** Description of Childhood Trauma Questionnaire

| | Item Description | Factors |
|---|---|---|
| Childhood Trauma Questionnaire: five factors | 1-I didn’t have enough to eat. | Physical Neglect |
| | 2-I was looked after and protected. | Physical Neglect |
| | 3-Someone in my family called me “stupid,” “lazy,” or “ugly.” | Emotional Abuse |
| | 4-My parents were too drunk or high to take care of the family. | Physical Neglect |
| | 5-I felt important or special in my family. | Emotional Neglect |
| | 6-I had to wear dirty or unsuitable clothes. | Physical Neglect |
| | 7-I felt loved. | Emotional Neglect |
| | 8-I thought my parents wished I had never been born. | Emotional Abuse |
| | 9-I got hurt so badly by someone in my family that I had to see a doctor. | Physical Abuse |
| | 10-There was nothing wrong with the way things were in my family. | - |
| | 11- Someone in my family hit me so hard that it left | Physical Abuse |

| | | |
|---|---|---|
| | bruises or marks. | |
| | 12- I was punished with a belt, a board, a cord, or some other hard object. | Physical Abuse |
| | 13-People in my family looked out for each other. | Emotional Neglect |
| | 14-Someone in my family said hurtful or insulting things to me. | Emotional Abuse |
| | 15-I believe that I was physically abused. | Physical Abuse |
| | 16-I had the perfect childhood. | - |
| | 17-I got hit or beaten so badly that it was noticed by someone like a teacher, neighbor, or doctor. | Physical Abuse |
| | 18-I felt hated by someone in my family. | Emotional Abuse |
| | 19-Members of my family felt close to each other. | Emotional Neglect |
| | 20-Someone tried to touch me in a sexual way or make me touch them. | Sexual Abuse |
| | 21-I was threatened to be made to do something sexual. | Sexual Abuse |
| | 22-I felt that my home was a good place to be. | - |
| | 23-Someone tried to make me do sexual things or watch sexual acts. | Sexual Abuse |
| | 24-Someone molested or acted sexually inappropriate with me. | Sexual Abuse |
| | 25-I was emotionally abused or tormented. | Emotional Abuse |
| | 26-Someone made sure I was healthy and taken care of. | Physical Neglect |
| | 27-I was sexually abused. | Sexual Abuse |
| | 28-My family was a source of strength and support. | Emotional Neglect |

Note: "-" indicates "not scored", as it is a validity item.

### Assessment of positive and negative natural environment

#### $PM_{2.5}$ concentration

Daily average $PM_{2.5}$ concentrations were obtained from the National Urban Air Quality Real-time Publishing Platform, which provides ground-level measurements for accurate exposure assessment. The Ozone Monitoring Instrument (OMI) offers long-term atmospheric species products at a spatial resolution of 13 km × 25 km. We used the L3-level OMI column products of atmospheric components from 2018 to 2025, retaining only data with a quality assurance value greater than 0.75. To achieve high-resolution $PM_{2.5}$ mapping with complete spatial coverage, we employed a machine-learning model to establish relationships between satellite products and ground observations, supported by auxiliary datasets. The Light Gradient Boosting Machine

(LightGBM) model was selected due to its established performance in remote sensing retrieval and atmospheric research[2,3]. The model is expressed as:

$$PM_{2.5}^{surface} = \text{LightGBM}(NDVI, t2, d2, sp, u10, v10, blh, e, RH, OMI)$$

Two evaluation metrics, $R^2$ and RMSE, were used to assess model performance through 10-fold cross-validation.

$$R^2 = 1 - \frac{\sum_{i-1}^{n}(y_{label}(i) - y_{mod}(i))^2}{\sum_{i-1}^{n}(y_{label}(i) - \overline{y_{mod}(i)})^2}$$

$$RMSE = \sqrt{\frac{1}{n} \times \sum_{i-1}^{n}(y_{label}(i) - y_{mod}(i))^2}$$

Where, $n$ is the total sample size, $y_{label}$ represents site-observed of $PM_{2.5}$ concentration, and $y_{mod}$ denotes model-estimated $PM_{2.5}$ concentration.

Both sample-based and spatial 10-fold cross-validations were applied. The dataset was randomly divided into ten subsets by sample and location. Nine subsets were used for training, and the remaining subset for testing. This process was repeated ten times, and model accuracy was evaluated across all test sets. The results demonstrated high accuracy of the $PM_{2.5}$ estimates.

***NDVI***

The *NDVI* is a widely used remote sensing metric for assessing vegetation health and density. It is calculated based on the reflectance of near-infrared (NIR) and red light:

$$\text{NDVI} = \frac{\text{NIR} - \text{RED}}{\text{NIR} + \text{RED}}$$

Values range from -1 to +1, with higher values indicating denser vegetation, and lower values (near or below zero) suggesting sparse vegetation, water bodies, or urban areas.

In this study, *NDVI* data were obtained from the MODIS 16-day global *NDVI* product (MOD13Q1), which provides consistent vegetation monitoring at a 250-meter spatial resolution. This dataset is particularly valuable for long-term ecological and environmental studies due to its global coverage and frequent updates (every 16 days).

**Extraction of exposure variables**

Residential addresses of all participants were geocoded using the Gaode Map API (https://lbs.amap.com/) to obtain corresponding longitude and latitude coordinates. The exposure window was defined as the five-year period preceding each participant's hospitalization or enrollment date. The corresponding $PM_{2.5}$ and *NDVI* values were then assigned to each participant's geocoded residential location. These environmental factors were linked to the patients' geographical information to construct an integrated patient disease-exposure database.

### MRI Data Acquisition and Preprocessing

**MRI Data Acquisition:** The T1W image were collected using a GE SIGNA 3T scanner (GE Healthcare) at the radiology department of Renmin Hospital of Wuhan University. The Sag 3D T1W parameters (Bravo) were set as follows: repetition time = 7.62 ms, echo time = 3.07 ms, flip angle = 12°, matrix = 256 × 256, field of view (FOV) = 256 × 256 $mm^2$, voxel size = 1 mm × 1 mm ×1 mm, slice gap = 0, and slices = 176. Participants with images of poor scan quality were excluded. Additionally, to check for between-group differences in image quality and head motion, the Euler number was computed for each T1W image[4].

**Preprocessing:** The T1W images underwent preprocessing in a surface-based space using the FreeSurfer software package (https://fsl.fmrib.ox.ac.uk/fsl/fslwiki). In short, the cortical surface was reconstructed through skull stripping, brain tissue segmentation, separation of cortical and subcortical regions, and creation of the interfaces and pial surfaces for white and gray matter. For each cortical region, surface area was extracted. To account for variations in brain size among participants, total intracranial volume (TIV) measurements were also obtained. We then regressed out the influences of sex, age and TIV.
